# Early Prognostic Instrumental and Laboratory Biomarkers in Post-MI

**DOI:** 10.1101/2023.05.13.23289438

**Authors:** Basheer Abdullah Marzoog, Ekaterina Vanichkina

## Abstract

**Background:** Post-myocardial infarction (MI) changes have been frequently reported in the literature and are associated with determining the prognosis.

**Aims:** To find a prognosis marker for the favorability of determination of the medium-term outcomes in patients with acute MI.

**Objectives:** MI patients’ prognosis is poorly understood and requires further elaboration.

**Materials and methods:** A single center, cross-sectional cohort study involved 211 patients’ medical history with acute MI, for the period 2014-2019, has been evaluated retrospectively for 76 parameters. The data collected from the Republic Rehabilitation Mordovian Hospital. The described measurement units are used in the local laboratories to describe the values. The descriptive values are expressed in the mean average and standard deviation. For statistical analysis, descriptive statistics, t-test independent by groups and dependent by numerical variables for repeated analysis for the same patients, multinomial logistic regression, Pearson’s correlation coefficient, ROC analysis, and for clarification purposes, diagrams and bar figures were used. For performing the statistical analysis, the SPSS program, version 28 used.

**Results:** Descriptive statistics showed a proportion of men to females 7:3. The mean age of the MI patients was 61.50 years (Std. Dev. ± 10.68), and the mean height of the sample was 171.00 cm (Std. Dev. ± 7.20). The mean body weight of the sample is 83.62 kg (Std. Dev. ± 12.35), and the body mass index (BMI) is 29.02 kg/m^2^ (Std. Dev. ± 5.07). The total hospitalization days are 14.79 (Std. Dev. ± 3.41). The mean heart rate (HR) beat per minute (bpm) was 79.03 (Std. Dev. ± 15.63), and the mean blood pressure was 138.53/84.09 mmHg (Std. Dev. ± 28.66/12.79). On the complete blood count (CBC), the mean level of the hemoglobin (Hb) 136.33 g/l (Std. Dev. ± 15.29), the mean level of the leukocytes (WBC) 8.76 /µl (Std. Dev. ± 2.77), the mean level of the red blood cells (RBC) 4.55 /µl (Std. Dev. ± 0.52), the mean level of the relative value of the lymphocytes 24.46 % (Std. Dev. ± 9.015), and the mean level of the thrombocytes 207.87 /µl (Std. Dev. ± 64.035). The mean erythrocytes segmentation rate (ESR) is 18.99 mm/hr (Std. Dev. ± 12.16). The regression analysis demonstrated that the dependent variable, complication, in particular, pericarditis, and the independent factor, concomitant disease, in particular, chronic heart failure, has a significant regression coefficient of 29.101 at p<0.05. Furthermore, the dependent variable, complication, in particular, pneumonitis, and the independent factor, concomitant disease, particularly, arrhythmia, have a significant regression coefficient of 21.937 at p<0.05.

**Conclusions:** An elevated level of CPK-MB/LDH/Troponin I is linked to the development of arrhythmia. Patients with other medical conditions experience high diastolic blood pressure and an enlargement of the right ventricle. The early complication observed after MI is the formation of a left ventricular aneurysm. Complications arise due to low levels of potassium and calcium. Chronic Kidney Disease (CKD) contributes to the End-Diastolic Size (EDS) of the Left Ventricle (LV), Troponin I, and creatine phosphokinase-MB (CPK-MB). Advanced CKD patients have a hypertrophic left ventricle and persistently elevated post-myocardial Infarction (MI) cardiac biomarkers (CPK-MB/LDH/Troponin I) due to impaired kidney detoxification. Therefore, prolonged elevation of MI biomarkers can be an indicative of severe MI or kidney function impairment due to the chronic mild elevation in the MI biomarkers. Pericarditis development is related to the pre-existence of chronic heart failure. Moreover, pneumonitis development is related to the pre-existence of arrhythmia.

**Others:** Hypertensive patients do not exhibit a significant increase in calcium levels, indicating that it is not a reliable biomarker in this patient population. Additionally, gender plays a crucial role in the development of ischemic heart disease, including myocardial infarction.

**Graphical abstract:** 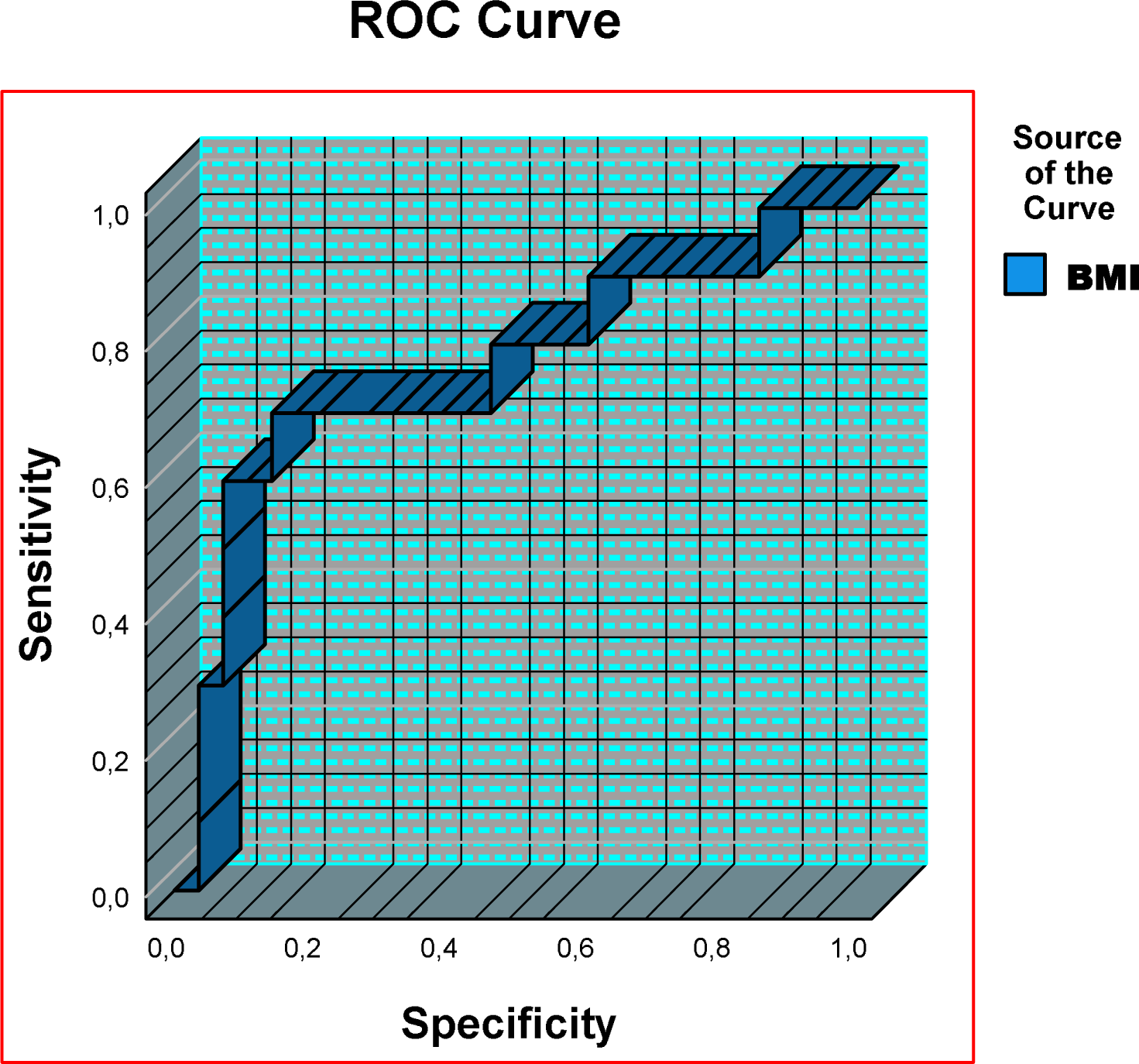

## Introduction

The heart regulates blood flow through intricate mechanisms involving the autonomic nervous and endocrine systems [1–4]. Disruptions in this delicate balance led to the development of both clinical and subclinical manifestations. Various factors, such as coronary vascular disease, can contribute to the decline of heart function. Unfortunately, patients with MI experience poor prognosis and quality of life due to the direct cardiovascular event or the related complication.

Current statistics indicate that cardiovascular events remain the leading cause of death and disability worldwide [5].

Many aspects of post-myocardial infarction (MI) changes remain unclear and require further investigation. Consequently, determining the precise life expectancy of MI patients remains challenging [6–8]. MI involves necrosis of myocardiocytes and disruption of the heart tissue’s essential functions, including automaticity, conduction, and contractility.

Several types of myocardial infarction exist including, transmural MI that involves full-heart wall thickness necrosis and falls under one of the following types of MI (MI type 1-5). According to the current European Society Of Cardiology guideline on the fourth definition of myocardial infarction, MI-1 is caused by rupture of the coronary atherosclerosis and subsequent thromboembolism, MI-2 is caused by coronary artery spasm (vasospasm), MI-3 is due to sudden cardiac death, MI-4a occurs after percutaneous coronary intervention (PCI), MI-4b occurs after coronary artery stenting, and MI-5 occurs after coronary artery bypass graft (CABG) [9,10].

A typical MI presentation includes distinct pain characteristics and localization that fail to respond to nitroglycerin, persists for over 15 minutes, and is accompanied by shortness of breath.

The pathophysiology of MI involves an imbalance between myocardial oxygen demand and supply [11,12]. Subsequently, cardiac cells experience ischemia, leading to sequelae such as oxidative stress due to mitochondrial and cellular membrane deterioration [7,13–23]. The lipid content of membranes plays a crucial role in cardiac cells regarding ischemia tolerance. Maintaining membrane stability during ischemia is cardioprotective [24].

Early prognostic instrumental and laboratory biomarkers in the post-MI period are used to assess the severity of myocardial damage and predict the likelihood of future cardiac events. Elevated levels of cardiac enzymes such as cardiac troponins, lactate dehydrogenase 1, and creatinine phosphokinase-MB fraction serve as a classical early laboratory indicators of myocardial cell necrosis. An electrocardiogram (ECG) is a rapid instrumental diagnostic method for MI. Measuring these biomarkers helps determine the extent of myocardial damage, identify complications, and develop a patient-tailored treatment plan. Early detection of these biomarkers is crucial for improving patient outcomes and reducing the risk of future cardiac events. However, these biomarkers alone are insufficient to assess systemic changes and damage to other organs, including the heart. Therefore, this study aimed to identify additional biomarkers that indicate early signs of damage to other organs (complications) in MI patients.

## Materials and Methods

This retrospective, single center, and observational study involved 211 patients diagnosed with myocardial infarction (MI) between 2014 and 2019. Primary data was collected from the Mordovian Republican Rehabilitation Hospital. (Figure 1) The analysis focuses on peak values of laboratory results obtained during the initial patient encounter.

**Figure 1.**
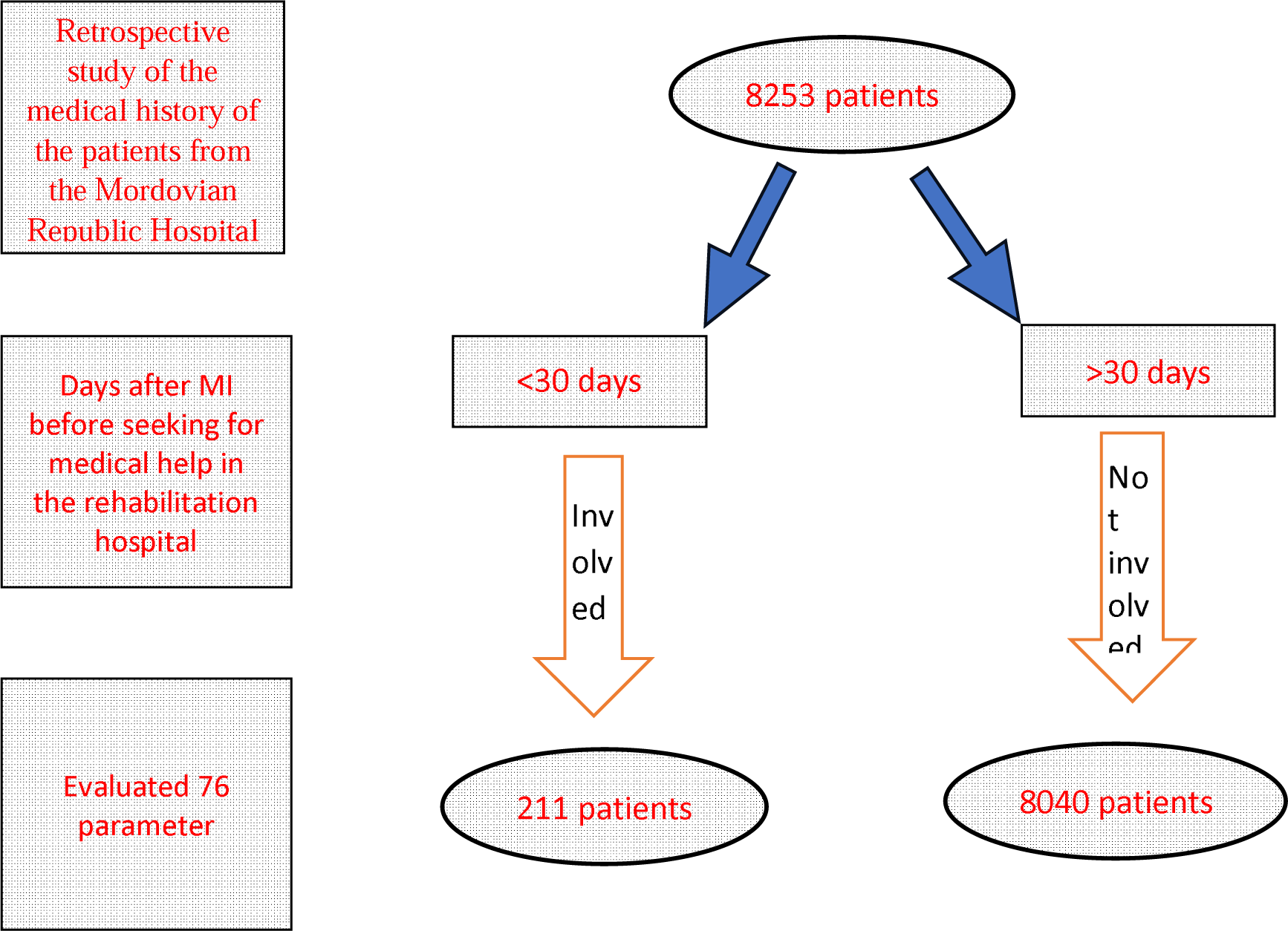
Diagrammatic presentation of the outflow of the study and sample selection. The data collected form the Mordovian Republic Rehabilitation Hospital for the period 2014-2019 (6 years).

Inclusion criteria involved seeking care at the rehabilitation hospital within 31 days of MI and participation in a rehabilitation program designed to improve post-MI prognosis and provide palliative care. Missing data was excluded from the analysis, and statistically significant values were identified using a p-value < 0.05. The Supplemental Material (Supplementary File 1) provides a detailed frequency table with absolute and relative values for each variable without missing data.

Measurement units used in the study were those of the local laboratory. The study protocol was approved by the Ethics Committee of the National Research Mordovia State University, Russia (approval number N8/2, dated 30.06.2022).

For statistical analysis, descriptive statistics, t-test independent by groups and dependent by numerical variables for repeated analysis for the same patients, multinomial logistic regression, Pearson’s correlation test, ROC analysis, and for clarification purposes used diagrams and bar figures. Not necessarily all the results of the mentioned statistical analysis exist in the paper because only the tests with statistically significant values have been described. For performing the statistical analysis, the SPSS program, version 28 used.

## Results

The descriptive statistics showed a proportion of male to female 7:3. The mean age of the MI patients was 61.50 years (Std. Dev. ± 10.68), and the mean height of the sample was 171.00 cm (Std. Dev. ± 7.20). The mean body weight of the sample is 83.62 kg (Std. Dev. ± 12.35), and the body mass index (BMI) is 29.02 kg/m^2^ (Std. Dev. ± 5.079). The total hospitalization days are 14.79 (Std. Dev. ± 3.41). Mean heart rate (HR) beat per minute (bpm) 79.03 (Std. Dev. ± 15.63) and mean blood pressure 138.53/84.09 mmHg (Std. Dev. ± 28.66/12.79). On the complete blood count (CBC), the mean level of the hemoglobin (Hb) 136.33 g/l (Std. Dev. ± 15.29), the mean level of the leukocytes (WBC) 8.76 /µl (Std. Dev. ± 2.77), the mean level of red blood cells (RBC) 4.55 /µl (Std. Dev. ± 0.52), the mean level of lymphocytes 24.469 % (Std. Dev. ± 9.01), and the mean level of the thrombocytes 207.87 /µl (Std. Dev. ± 64.03). The mean segmentation rate (ESR) is 18.99 mm/hour (Std. Dev. ± 12.16).

In terms of biochemical analysis, the mean level of alanine transpeptidase (ALT) is 41.12 U/l (Std. Dev. ± 42.97), and the mean level of the aspartate transpeptidase (AST) is 58.78 U/l (Std. Dev. ± 71.11). Furthermore, the mean level of creatinine is 82.76 µmol/l (Std. Dev. ± 29.52), the mean level of urea is 6.694 mmol/l (Std. Dev. ± 2.07), and the mean level of venous glucose 6.997 mmol/l (Std. Dev. ± 2.98). The mean level of the total bilirubin is 16.204 mmol/l (Std. Dev. ± 6.45).

Changes in lipid profile are observed; the mean total cholesterol level is 5.27 mmol/l (Std. Dev. ± 1.33), low-density lipoprotein (LDL) 3.25 mmol/l (Std. Dev. ± 1.09), and the triacylglycerol (TAG) 1.72 mmol/l (Std. Dev. ± 0.91). Additionally, changes in the coagulogram results are also observed: the mean level of the activated partial thromboplastin time (aPTT) 27.82 s (Std. Dev. ± 8.09), and the mean value of international normalization ratio (INR) 1.10 (Std. Dev. ± 0.37).

Electrolyte changes are expressed in MI patients, with the mean sodium level in serum 142.241 mmol/l (Std. Dev. ± 4.83), and the mean level of potassium 4.561 mmol/l (Std. Dev. ± 6.64), and the mean calcium level 1.283 mmol/l (Std. Dev. ± 1.06).

Specific cardiac biomarker changes are presented in the troponin I and creatinine phosphokinase-MB fraction and lactate dehydrogenase 1. The mean level of troponin-I was 5.34 ng/l (Std. Dev. ± 8.64), and the mean level of the creatinine phosphokinase-MB fraction (CPK-MB) 100.364 U/l (Std. Dev. ± 259.36). Furthermore, the mean lactate dehydrogenase-1 (LDH-1) level was 685.59 U/l (Std. Dev. ± 574.20). The mean level of myoglobin is 111.30 ng/ml (Std. Dev. ± 76.83).

Changes in the instrumental findings, including echocardiogram changes. The mean diameter of the aorta at the level of the Valsalva sinus is 35.57 mm (Std. Dev. ± 4.30). The mean ejection fraction (EF) is 54.19 % (Std. Dev. ± 9.45). The mean end-diastolic volume (EDV) is 138.74 ml (Std. Dev. ± 39.18). Additionally, the mean end-diastolic size of the left ventricle, left atrium, right ventricle, and right atrium 52.87 mm, 40.29 mm, 26.93 mm, 36.06 mm (Std. Dev. ± 7.47/5.15/ 4.07/ 4.45), respectively.

In the general urine analysis, the mean level of urine protein was 1.83 (Std. Dev. ± 11.82), and the mean level of the urine WBC in the field of vision was 2.80 (Std. Dev. ± 7.87). And the mean level of epithelial cells in vision is 1.61 (Std. Dev. ± 2.45).

In terms of pre-MI concomitant disease, arterial hypertension stands in the first place. (*Figure 2*)

**Figure 2.**
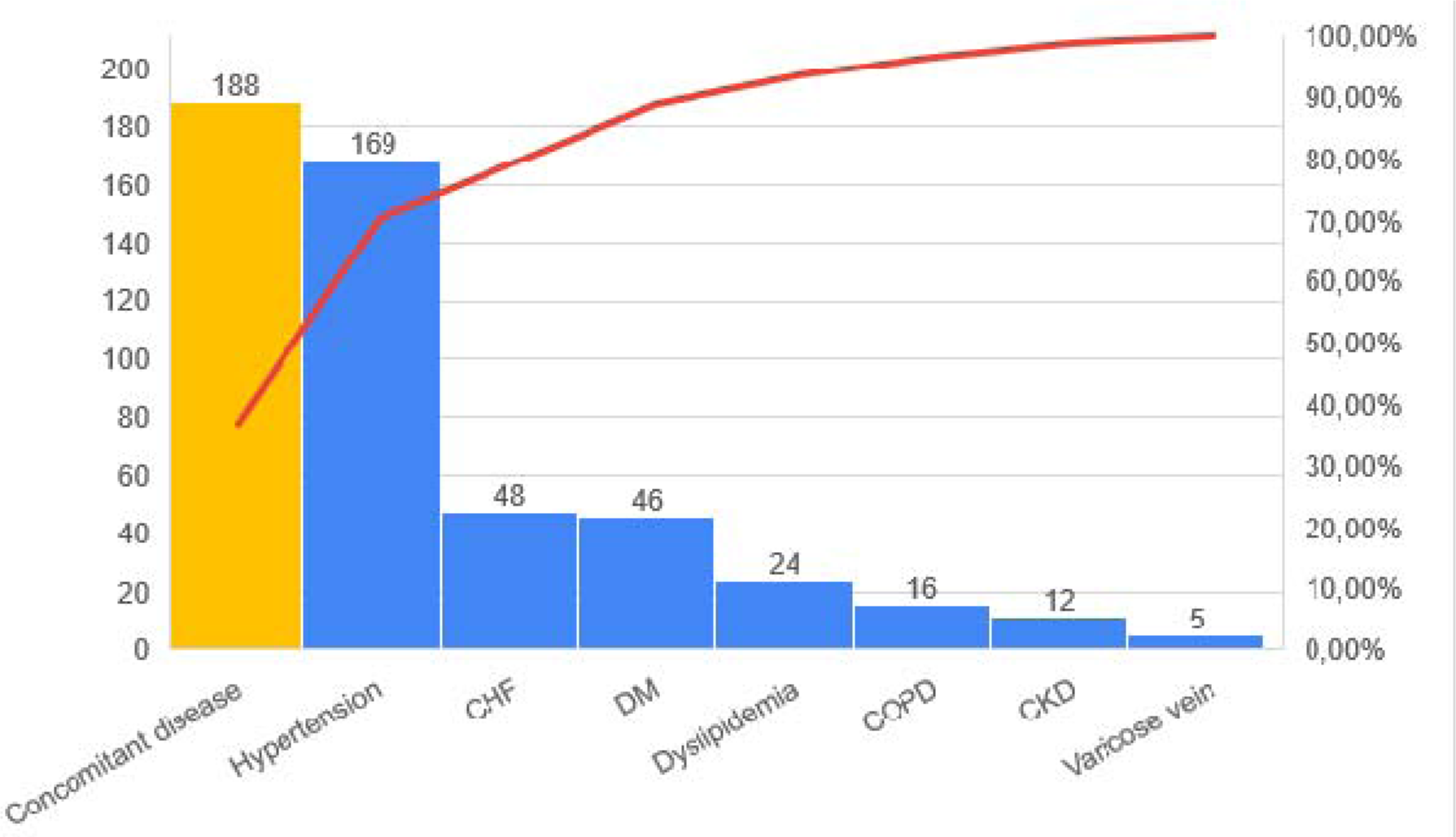
Most frequently concomitant diseases reported in MI patients. Descending arranges for the concomitant diseases by frequency of occurrence. Concomitant disease exists in 188 (>90%) patients out of 211. Abbreviations: CHF; chronic heart failure, DM; diabetes mellitus, COPD; chronic obstructive lung disease, CKD; chronic kidney disease.

The most frequently reported post-MI complication is heart failure and various types of arrhythmia. In patients with pre-existing CHF, the degree or severity of heart failure has worsened and changed to a more advanced stage. (*Figure 3*)

**Figure 3.**
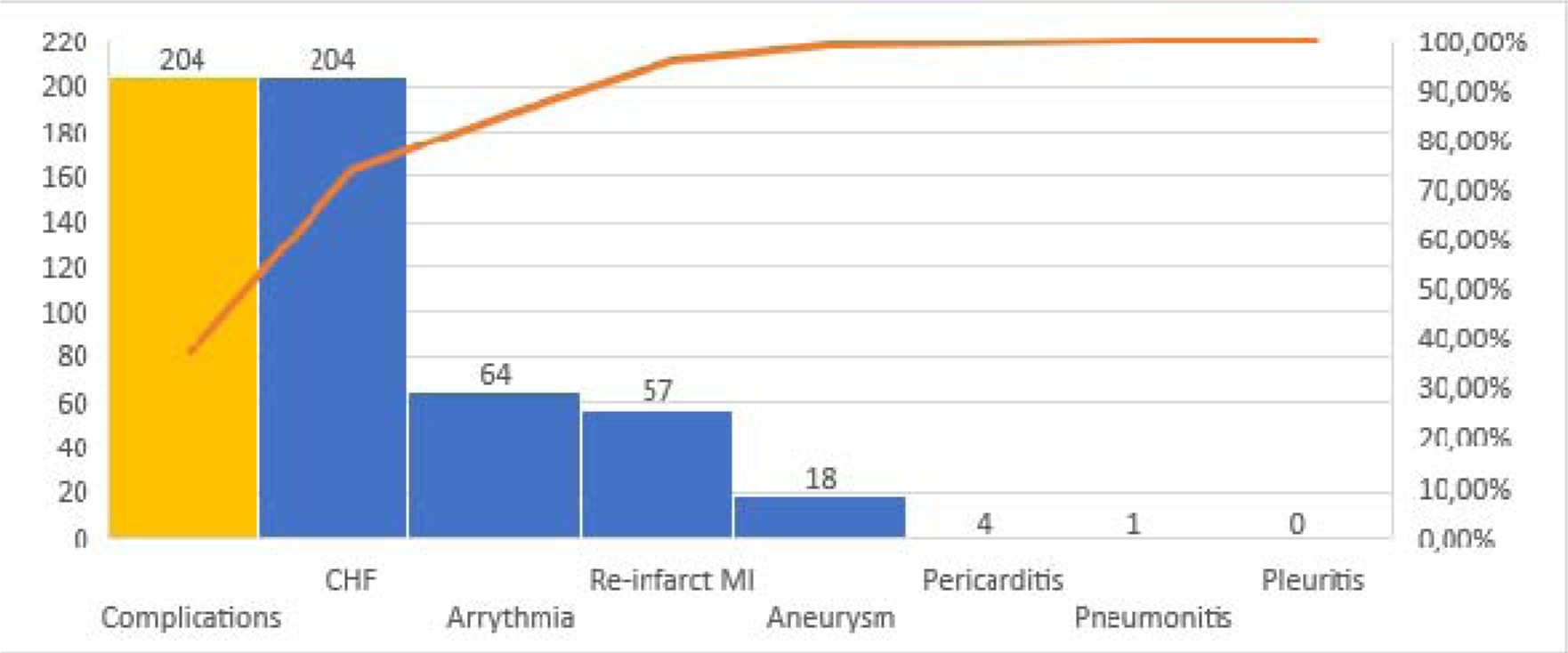
Most frequently reported complications post PCI and MI in patients. Descending arranges for the complications by frequency of occurrence. Complications exist in 204 (>90%) patients out of 211. Abbreviations: CHF; chronic heart failure, MI; myocardial
infarction.

Several factors contribute to the post-MI prognosis worsening, as shown in Table 1. The variability in these parameters is associated with the grouping factors. The cut age at which females develop MI is ≥ 73 years old.

**Table 1:** Results of the T-test to group the sample according to different principles to assess the potential role of each. Only statically significant parameters have been shown in the table.

|  |  |
| --- | --- |
| Variable | T-tests; Grouping: Gender<br>Group 1: F |

|  | Group 2: M |  |  |  |  |
| --- | --- | --- | --- | --- | --- |
|  | Mean<br>F | Mean<br>M | t-value | df | p |
| Age | 65.26 | 59.73 | 3.72539 | 209 | 0.000251 |
| Height | 164.67 | 173.07 | -3.84356 | 35 | 0.000490 |
| BMI | 31.96 | 27.81 | 2.50249 | 36 | 0.017012 |
| SBP | 144.40 | 135.41 | 2.25373 | 205 | 0.025272 |
| Aorta diameter at Valsalva sinus | 33.19 | 36.68 | -5.78550 | 200 | 0.000000 |
| EDV (ML) | 128.60 | 143.56 | -2.36302 | 169 | 0.019264 |
| EDS of RA (MM) | 35.06 | 36.53 | -2.17269 | 193 | 0.031022 |
| Hb (g/l) | 129.89 | 139.39 | -4.29775 | 200 | 0.000027 |
| RBC /L | 4.44 | 4.62 | -2.16953 | 163 | 0.031491 |
| Urine epithelial cells | 2.41 | 1.25 | 3.12240 | 191 | 0.002073 |
| Myoglobin (ng/ml) | 199.90 | 67.00 | 19.18246 | 1 | 0.033158 |
| LDH 2 (U/l) | 409.91 | 713.16 | -2.47399 | 71 | 0.015754 |
| Glucose (mmol/l) | 7.79 | 6.63 | 2.53778 | 192 | 0.011950 |

| Variable | T-tests; Grouping: Arrhythmia |  |  |  |  |
| --- | --- | --- | --- | --- | --- |
|  | Group 1: Yes |  |  |  |  |
|  | Group 2: No |  |  |  |  |
|  | Mean<br>Yes | Mean<br>No | t-value | df | p |
| Total hospitalization days | 15.53 | 14.56 | 2.07859 | 205 | 0.038900 |
| Age | 63.89 | 60.46 | 2.23887 | 205 | 0.026239 |
| EF (%) | 50.85 | 55.50 | -3.32859 | 196 | 0.001043 |
| EDS of LA (MM) | 42.05 | 39.53 | 3.24848 | 196 | 0.001365 |
| EDS of RA (MM) | 37.88 | 35.24 | 3.92822 | 192 | 0.000119 |
| Troponin I (ng/ml) | 8.36 | 4.00 | 3.20317 | 176 | 0.001614 |
| CPK-MB (U/l) | 172.83 | 69.12 | 2.50073 | 178 | 0.013297 |
| LDH (U/l) | 906.96 | 589.07 | 3.17184 | 143 | 0.001854 |
| ALT (mmol/l) | 54.25 | 35.44 | 2.86748 | 193 | 0.004597 |
| AST (mmol/l) | 78.37 | 48.51 | 2.81321 | 194 | 0.005410 |
| TAG (mmol/l) | 1.50 | 1.83 | -2.34537 | 191 | 0.020034 |

| Variable | T-tests; Grouping: DM |  |  |  |  |
| --- | --- | --- | --- | --- | --- |
|  | Group 1: No |  |  |  |  |
|  | Group 2: Yes |  |  |  |  |
|  | Mean<br>No | Mean<br>Yes | t-value | df | p |
| Age | 60.55 | 64.96 | -2.5331 | 204 | 0.012057 |
| Aorta diameter at Valsalva sinus | 35.87 | 34.39 | 2.0399 | 196 | 0.042697 |
| EDS of LV (MM) | 52.20 | 54.87 | -2.0736 | 181 | 0.039529 |
| Hb (g/l) | 137.75 | 131.91 | 2.2609 | 195 | 0.024871 |
| Myoglobin (ng/ml) | 67.00 | 199.90 | -19.1825 | 1 | 0.033158 |
| TAG (mmol/l) | 1.62 | 2.14 | -3.3127 | 189 | 0.001107 |
| Glucose (mmol/l) | 6.17 | 9.90 | -8.3336 | 188 | 0.000000 |

| Variable | T-tests; Grouping: Hypertension |  |  |  |  |
| --- | --- | --- | --- | --- | --- |
|  | Group 1: Yes |  |  |  |  |
|  | Group 2: No |  |  |  |  |
|  | Mean<br>Yes | Mean<br>No | t-value | df | p |
| Age | 62.24 | 58.32 | 2.06235 | 204 | 0.040442 |
| SBP | 140.73 | 128.33 | 2.50247 | 200 | 0.013134 |
| DBP | 85.15 | 80.14 | 2.25165 | 199 | 0.025437 |
| EDS of LA (MM) | 40.68 | 38.45 | 2.27065 | 195 | 0.024261 |
| LDH 1 (U/l) | 643.44 | 921.24 | -2.21515 | 140 | 0.028365 |
| INR | 1.05 | 1.35 | -4.21313 | 174 | 0.000040 |
| TAG (mmol/l) | 1.79 | 1.44 | 2.01466 | 189 | 0.045358 |
| Ca (mmol/l) | 1.20 | 1.68 | -2.18942 | 162 | 0.029997 |

| Variable | T-tests; Grouping: CKD |  |  |  |  |
| --- | --- | --- | --- | --- | --- |
|  | Group 1: No |  |  |  |  |
|  | Group 2: Yes |  |  |  |  |
|  | Mean | Mean | t-value | df | p |
|  | No | Yes |  |  |  |
| EDS of LV (MM) | 52.49 | 57.50 | -2.16124 | 178 | 0.032013 |
| Troponin I (ng/ml) | 4.77 | 19.25 | -4.85863 | 172 | 0.000003 |
| CPK-MB 1(U/l) | 89.05 | 372.14 | -3.20560 | 171 | 0.001609 |

| Variable | T-tests; Grouping: COPD |  |  |  |  |
| --- | --- | --- | --- | --- | --- |
|  | Group 1: No |  |  |  |  |
|  | Group 2: Yes |  |  |  |  |
|  | Mean | Mean | t-value | df | p |
|  | No | Yes |  |  |  |
| EDS of RV (MM) | 26.81 | 28.91 | -1.99714 | 195 | 0.047201 |
| leukocytes /L | 8.64 | 10.08 | -2.02454 | 193 | 0.044292 |
| Urine protein | 1.27 | 8.90 | -2.41397 | 191 | 0.016724 |
| LDH 1 (U/l) | 653.47 | 1047.86 | -2.47174 | 140 | 0.014645 |
| ALT (mmol/l) | 38.93 | 63.81 | -2.22692 | 192 | 0.027115 |

| Variable | T-tests; Grouping: Dyslipidemia |  |  |  |  |
| --- | --- | --- | --- | --- | --- |
|  | Group 1: No |  |  |  |  |
|  | Group 2: Yes |  |  |  |  |
|  | Mean | Mean | t-value | df | p |
|  | No | Yes |  |  |  |
| Total cholesterol (mmol/l) | 5.21 | 5.92 | -2.45779 | 191 | 0.014871 |
| LDL (mmol/l) | 3.20 | 3.77 | -2.42518 | 187 | 0.016251 |
| Na (mmol/l) | 142.50 | 140.01 | 2.17050 | 174 | 0.031324 |
| Ca (mmol/l) | 1.34 | 0.81 | 1.99261 | 162 | 0.047984 |

| Variable | T-tests; Grouping: Varicose vein |  |  |  |  |
| --- | --- | --- | --- | --- | --- |
|  | Group 1: No<br>Group 2: Yes |  |  |  |  |
|  | Mean<br>No | Mean<br>Yes | t-value | df | p |
| ESR (mm/hr.) | 18.28 | 33.40 | -2.86024 | 190 | 0.004707 |
| Fibrinogen (mg/dl) | 3439.63 | 4325.00 | -2.39330 | 162 | 0.017842 |

| Variable | T-tests; Grouping: Complications |  |  |  |  |
| --- | --- | --- | --- | --- | --- |
|  | Group 1: Yes<br>Group 2: No |  |  |  |  |
|  | Mean<br>Yes | Mean<br>No | t-value | df | p |
| Total hospitalization days | 14.91 | 11.43 | 2.89231 | 207 | 0.004233 |
| EDS of RA (MM) | 35.91 | 40.40 | -2.25749 | 192 | 0.025103 |
| K (mmol/l) | 4.09 | 21.53 | -6.37437 | 182 | 0.000000 |
| Ca (mmol/l) | 1.20 | 4.07 | -6.66743 | 166 | 0.000000 |

| Variable | T-tests; Grouping: Aneurysm |  |  |  |  |
| --- | --- | --- | --- | --- | --- |
|  | Group 1: No<br>Group 2: Yes |  |  |  |  |
|  | Mean<br>No | Mean<br>Yes | t-value | df | p |
| EF (%) | 55.18 | 43.59 | 5.13333 | 198 | 0.000001 |
| Urine epithelial cells | 1.44 | 3.63 | -3.51931 | 191 | 0.000541 |
| Troponin I (ng/ml) | 4.99 | 9.99 | -2.02501 | 178 | 0.044361 |

| Variable | T-tests; Grouping: Pneumonitis |  |  |  |  |
| --- | --- | --- | --- | --- | --- |
|  | Group 1: No |  |  |  |  |
|  | Group 2: Yes |  |  |  |  |
|  | Mean<br>No | Mean<br>Yes | t-value | df | p |
| Aorta diameter at Valsalva sinus | 35.66 | 20.00 | 3.73889 | 199 | 0.000242 |
| leukocytes /L | 8.72 | 14.80 | -2.20307 | 197 | 0.028749 |
| ESR (mm/hr.) | 18.78 | 48.00 | -2.42454 | 194 | 0.016244 |
| Urine leukocytes | 2.72 | 22.00 | -2.46896 | 197 | 0.014402 |
| CPK-MB (U/l) | 94.87 | 1164.00 | -4.29558 | 178 | 0.000029 |

| Variable | T-tests; Grouping: D-dimer (ng/l) |  |  |  |  |
| --- | --- | --- | --- | --- | --- |
|  | Group 1: Increased |  |  |  |  |
|  | Group 2: Normal |  |  |  |  |
|  | Mean<br>Increased | Mean<br>Normal | t-value | df | p |
| LDH 1 (U/l) | 353.88 | 808.50 | -2.28113 | 10 | 0.045698 |

| Variable | T-tests; Grouping: Pericarditis |  |  |  |  |
| --- | --- | --- | --- | --- | --- |
|  | Group 1: No |  |  |  |  |
|  | Group 2: Yes |  |  |  |  |
|  | Mean<br>No | Mean<br>Yes | t-value | df | p |
| EDS of LA (MM) | 40.19 | 45.50 | -2.05662 | 199 | 0.041028 |

| Variable | T-tests; Grouping: Re-infarct MI |  |  |  |  |
| --- | --- | --- | --- | --- | --- |
|  | Group 1: Yes |  |  |  |  |
|  | Group 2: No |  |  |  |  |
|  | Mean<br>Yes | Mean<br>No | t-value | df | p |
| Age | 64.07 | 60.75 | 2.08949 | 208 | 0.037879 |

Several intriguing correlations were described in this study, such as a proportional relationship between the HR and EF, as depicted in (*Figure 4*). Additional correlations are presented in Table 2; further visual representations can be found in Supplementary File 2.

**Figure 4.**
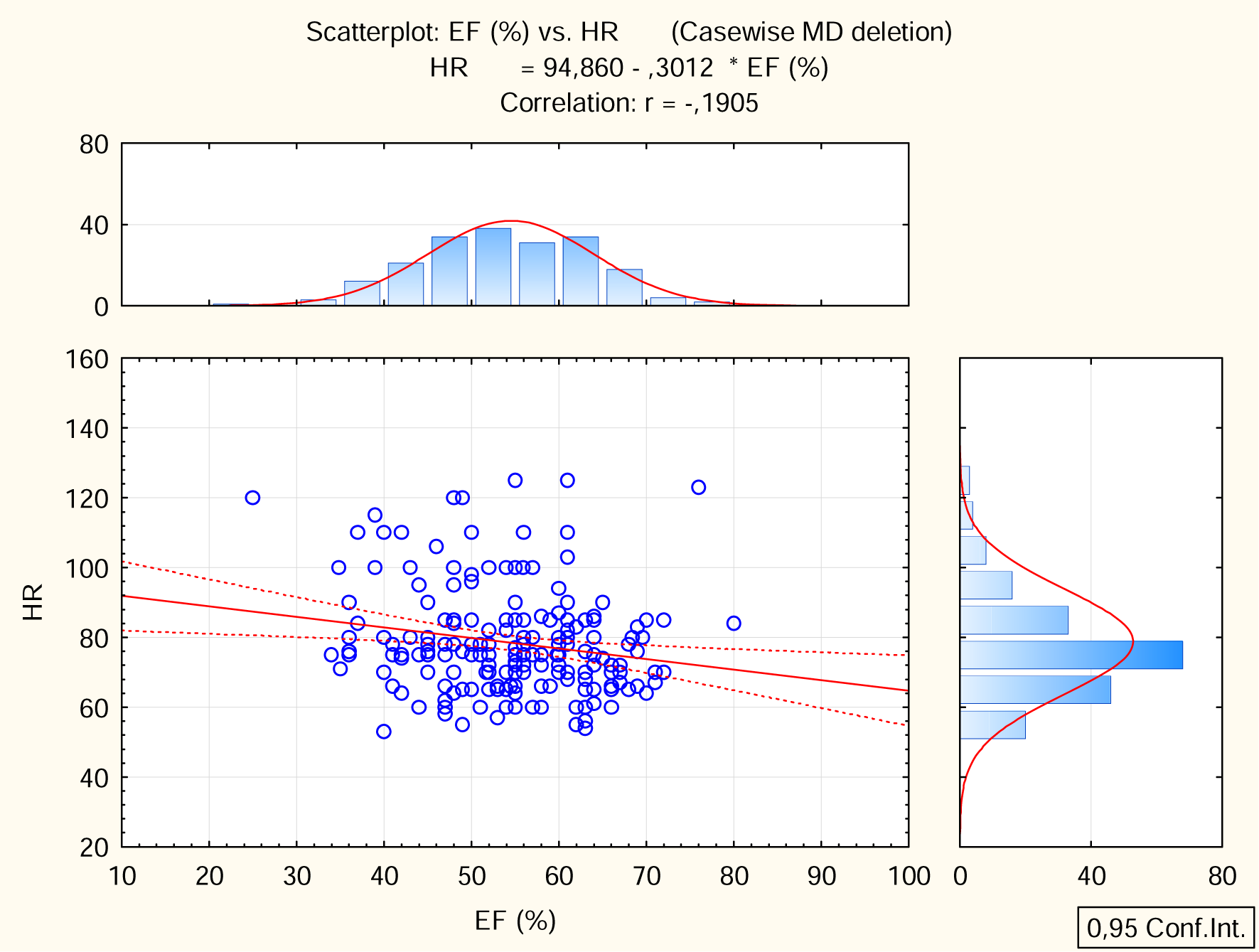
Proportional correlation between the heart rate and the ejection fraction.

The regression analysis demonstrated that the dependent variable, complication, in particular, pericarditis, and the independent factor, concomitant disease, in particular, chronic heart failure, has a significant regression coefficient of 29.101 at p<0.05. Furthermore, the dependent variable, complication, in particular, pneumonitis, and the independent factor, concomitant disease, particularly, arrhythmia, have a significant regression coefficient of 21.937 at p<0.05. (Table 3A, 3B)

**Table 3:**
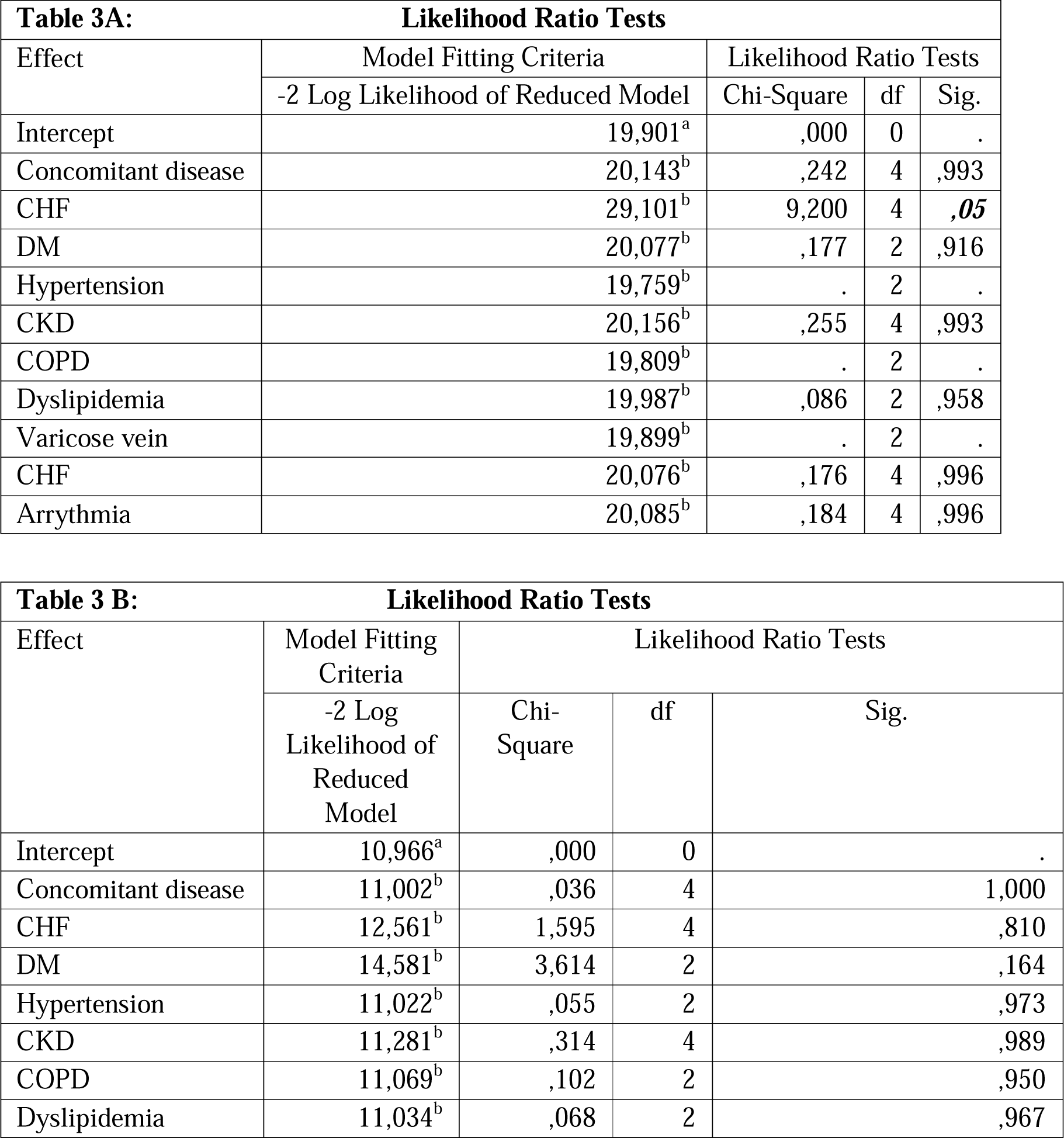
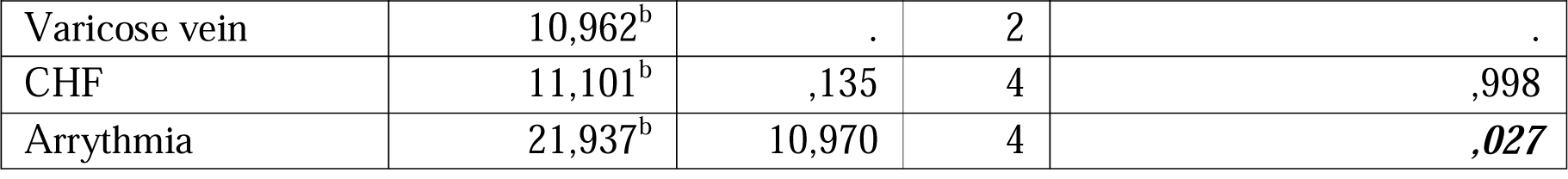
Multinomial logistic regression A-Pericarditis as a dependent variable on the pre-existence of chronic heart failure. B-Pneumonitis as a dependent variable on the pre-existence of arrythmia.

| **Table 3A: Likelihood Ratio Tests** |  |  |  |  |
| --- | --- | --- | --- | --- |
| Effect | Model Fitting Criteria | Likelihood Ratio Tests |  |  |
|  | -2 Log Likelihood of Reduced Model | Chi-Square | df | Sig. |
| Intercept | 19,901<sup>a</sup> | ,000 | 0 | . |
| Concomitant disease | 20,143<sup>b</sup> | ,242 | 4 | ,993 |
| CHF | 29,101<sup>b</sup> | 9,200 | 4 | **,05** |
| DM | 20,077<sup>b</sup> | ,177 | 2 | ,916 |
| Hypertension | 19,759<sup>b</sup> | . | 2 | . |
| CKD | 20,156<sup>b</sup> | ,255 | 4 | ,993 |
| COPD | 19,809<sup>b</sup> | . | 2 | . |
| Dyslipidemia | 19,987<sup>b</sup> | ,086 | 2 | ,958 |
| Varicose vein | 19,899<sup>b</sup> | . | 2 | . |
| CHF | 20,076<sup>b</sup> | ,176 | 4 | ,996 |
| Arrhythmia | 20,085<sup>b</sup> | ,184 | 4 | ,996 |

| **Table 3 B: Likelihood Ratio Tests** |  |  |  |  |
| --- | --- | --- | --- | --- |
| Effect | Model Fitting Criteria | Likelihood Ratio Tests |  |  |
|  | -2 Log Likelihood of Reduced Model | Chi-Square | df | Sig. |
| Intercept | 10,966<sup>a</sup> | ,000 | 0 | . |
| Concomitant disease | 11,002<sup>b</sup> | ,036 | 4 | 1,000 |
| CHF | 12,561<sup>b</sup> | 1,595 | 4 | ,810 |
| DM | 14,581<sup>b</sup> | 3,614 | 2 | ,164 |
| Hypertension | 11,022<sup>b</sup> | ,055 | 2 | ,973 |
| CKD | 11,281<sup>b</sup> | ,314 | 4 | ,989 |
| COPD | 11,069<sup>b</sup> | ,102 | 2 | ,950 |
| Dyslipidemia | 11,034<sup>b</sup> | ,068 | 2 | ,967 |
| Varicose vein | 10,962<sup>b</sup> | . | 2 | . |
| CHF | 11,101<sup>b</sup> | ,135 | 4 | ,998 |
| Arrhythmia | 21,937<sup>b</sup> | 10,970 | 4 | **,027** |

The chi-square statistic is the difference in −2 log-likelihoods between the final model and a reduced model. The reduced model is formed by omitting an effect from the final model. The null hypothesis is that all parameters of that effect are 0. Value in bold and italic is statically significant. a. This reduced model is equivalent to the final model because omitting the effect does not increase the degrees of freedom. b. Unexpected singularities in the Hessian matrix are encountered. This indicates that either some predictor variables should be excluded or some categories should be merged.

Additionally, results of the Receiver Operating Characteristic (ROC) analysis presented in a separate supplementary file. (Supplementary file 3)

## Discussion

Elevated blood pressure and being overweight are significant risk factors for the development of myocardial infarction (MI), especially for women at early menopausal age [25–27]. Gender also plays a role in the occurrence of ischemic heart disease, with males having higher systolic blood pressure and a larger aortic diameter at the Valsalva sinus [27].

Patients with other medical conditions often experience high diastolic blood pressure, leading to heart failure or the calcification of elastic and musculoelastic blood vessels [28,29]. High diastolic blood pressure, or diastolic hypertension, is a diastolic blood pressure reading above 80 mm Hg in individuals with normal systolic blood pressure [28]. Increases in diastolic pressure in individuals between the ages of 40 and 89 years double the risk of heart disease or stroke for every 10 mm Hg increase in diastolic pressure [28]. Obesity, high triglyceride levels, smoking, and alcohol consumption are potential contributors to high diastolic blood pressure [28].

Additionally, patients with concomitant diseases tend to have an enlarged right ventricle, indicating that heart failure is usually on the right side of the heart [30]. Right ventricular hypertrophy can occur in response to pressure overload, most commonly due to severe lung disease [30]. Symptoms of right ventricular hypertrophy may include exertional chest pain, peripheral edema, exertional syncope, and right upper quadrant pain [30].

Elderly patients with concomitant diabetes mellitus often have a smaller aortic diameter and left ventricular size [31]. A case-control study found that abdominal aortic diameter was larger in diabetic patients with peripheral arterial disease than in healthy controls [31]. However, further research is needed to understand the exact relationship between abdominal aortic diameter and diabetes.

Diabetic patients with low levels of hemoglobin (Hb) and high levels of triglycerides (TAG) may experience a significant increase in post-myocardial infarction (MI) myoglobin [31]. Myoglobin is a protein found in muscle tissue, and its levels can increase after a heart attack [31].

Hypertensive patients, who are typically of advanced age, have isolated systolic blood pressure, which tends to be high [32]. Post-MI hypertensive patients have low levels of calcium (Ca+2), international normalized ratio (INR), and lactate dehydrogenase (LDH), as well as high levels of TAG and extravascular fluid in the left atrium [32]. This indicates that high calcium levels may not be a reliable biomarker for hypertensive patients [32]. Hypertensive patients also do not exhibit high coagulation time or LDH levels after MI compared to non-hypertensive patients [21,32]. Hypertensive patients have a hypertrophic left atrium and hypertriglyceridemia, although dyslipidemia may be more typical in hypertensive patients [32].

Chronic Kidney Disease (CKD) contributes to the End-Diastolic Size (EDS) of the Left Ventricle (LV), Troponin I, and creatine phosphokinase-MB (CPK-MB). Advanced CKD patients often have a hypertrophic left ventricle and persistently elevated post-myocardial Infarction (MI) cardiac biomarkers due to impaired kidney detoxification [33,34].

Chronic Obstructive Pulmonary Disease (COPD) is associated with right ventricle hypertrophy and increased EDS of the Right Ventricle (RV). COPD patients often have high serum leukocytes, proteinuria, serum Lactate Dehydrogenase (LDH), and serum Alanine Aminotransferase (ALT), indicating chronic inflammation and prolonged liver intoxication [35].

MI patients often have high total cholesterol, Low-Density Lipoprotein (LDL), and low calcium and sodium levels, supporting the hypothesis that dyslipidemia is a significant risk factor for MI [36].

Post-MI complications can lead to prolonged hospitalization in the Intensive Care Unit (ICU). These complications are associated with low EDS of the Right Atrium (RA), hypokalemia, and hypocalcemia. Arrhythmia is a common post-MI complication, often leading to prolonged ICU stays and elevated levels of Left Atrium (LA) EDS, RA troponin I, CPK-MB, LDH, ALT, and Aspartate Aminotransferase (AST) [37,38].

Reinfarction is common in older age. Early complications include a left ventricular aneurysm associated with elevated urine epithelial cells and troponin I. Late post-MI complications include Dressler syndrome, which includes pericarditis, pneumonitis, and pleuritis [39–41].

MI patients often exhibit laboratory indicators of a chronic inflammatory process, such as an elevated Erythrocyte Sedimentation Rate (ESR) level. High AST is also a characteristic feature of patients in the acute stage of MI. MI patients may experience heart failure with preserved ejection fraction (HFpEF) [42,43].

A single study has suggested that elevated troponin T may be possible in patients with neuromuscular diseases [44]. Therefore, a separate elevation of cardiac troponin T is insufficient for diagnosing MI [36].

Regarding post-MI laboratory changes, COPD patients exhibit significantly elevated levels of LDH. In patients with concomitant hypertension after MI, there is a decrease in Ca+2, INR, and LDH levels and an increase in TAG and EDS of the left atrium. Dyslipidemia is more commonly observed in hypertensive patients, who also exhibit hypertriglyceridemia and left atrium hypertrophy. Patients with concomitant CKD experience worsening EDS of LV, troponin I, and CPK-MB levels. In diabetic patients, there is a decrease in Hb levels, an increase in TAG levels, and a significant elevation in post-MI myoglobin levels [45].

Early complications include a left ventricle aneurysm that is seen with elevated urine epithelial cells and troponin I. Reinfarction is found to be reported in older age as a nonmodified risk factor. MI patients have a laboratory indicator of the chronic inflammatory process presented by elevation of ESR level. High AST is a characteristic feature of patients with MI in the acute stage. Early post-MI patients have preserved EF heart failure (HFpEF). Interestingly, MI complicated with pneumonitis is associated with higher plasma leukocytes, ESR, Urine leukocytes, and CPK-MB. Post-MI remodeling is a classic change supported by most studies [46].

The high CPK-MB/LDH/Troponin I is associated with arrhythmia development. Suggesting that a high level of any of the cardiac biomarkers is an indicator of arrhythmia development [20].

Regarding novel nonclassical risk factors for cardiovascular disease development, Basheer Marzoog conducted a clinical trial using two mass spectrometers to demonstrate the changes in the exhaled breath analysis in patients with ischemic heart disease. The clinical trial in the recruiting phase. Several risk factors can cause cardiac damage, including infectious factors such as COVID-19 [13].

Regarding the molecular biopathophysiology of myocardial infarction suggested to involve lipid peroxidation changes and further autophagy dysregulation. Lipid peroxidation plays a significant role in myocardiocyte necrosis, serving as a critical step in its pathogenesis. Additionally, under ischemic conditions, myocardiocytes naturally upregulate the of the autophagy as a protective mechanism, aiming to salvage themselves from ischemic damage by self-digesting impaired organelles and intracellular components. However, in prolonged ischemia, as in MI, autophagy biomarkers levels significantly elevate as the ischemic stage progresses. Therefore, autophagy biomarkers can serve as an indicator of myocardial cell ischemia, particularly in cases of short-term ischemia. Monitoring the progression of ischemia by assessing serum autophagy biomarker levels is a promising approach [16].

Currently, scientists face the challenge of developing of disease-related autophagy biomarkers. One potential solution is exposing myocardiocytes to ischemia in vitro and evaluating specific autophagy biomarkers that are selective for myocardiocytes. This approach could enable the utilization of autophagy biomarkers as an early diagnostic tool for pre-myocardial infarction (ischemic period) to prevent myocardiocyte necrosis.

## Conclusions

Gender plays an essential role in the development of ischemic heart disease, including MI [6,47–50]. A proportional correlation between HR and EF. Patients with concomitant diseases have high diastolic blood pressure, which is related to heart failure and or the calcification of the elastic and musculoelastic blood vessels. Patients with concomitant diseases have a larger right ventricle size. They suggest that heart failure is usually on the right side of the heart (large circulation).

Pericarditis development is related to the pre-existence of chronic heart failure. Moreover, pneumonitis development is related to the pre-existence of arrhythmia.

Hypertensive patients do not exhibit a significant increase in calcium levels, indicating that it is not a reliable biomarker in this patient population. Additionally, gender plays a crucial role in the development of ischemic heart disease, including myocardial infarction.

This study is limited by its retrospective design, which introduces several potential sources of bias. One major limitation is the inability to determine the survival rate and hazard ratio, as it is difficult to ascertain which patients developed additional complications or ultimately passed away. Additionally, since retrospective studies rely on existing data, there is limited control over data quality, which may need to be completed or updated. Selection bias is also a risk, as the study sample may differ from the studied population. Furthermore, controlling for confounding variables in retrospective studies is difficult, which may affect the results. Finally, the limited ability to generalize findings is another potential limitation, as retrospective studies may not reflect real-world conditions or be generalizable to other populations or settings.

## List of abbreviations

CHF: chronic heart failure
DM: Diabetes mellitus
CKD: Chronic kidney disease
COPD: Chronic obstructive pulmonary disease
MI: myocardial infarction
HR: heart rate
ESR: erythrocytes sedimentation rate
RBC: red blood cells

## Declarations

### Ethics approval and consent to participate

The study approved by the National Research Mordovia State University, Russia, from “Ethics Committee Requirement N8/2 from 30.06.2022”.

### Consent for publication

Written informed consent was obtained from the participants for publication of study results and any accompanying images.

### Availability of data and materials

applicable on reasonable request.

### Competing interests

The authors declare that they have no competing interests regarding publication.

### Funding’s

The work of **Basheer Abdullah Marzoog** was financed by the Ministry of Science and Higher Education of the Russian Federation within the framework of state support for the creation and development of World-Class Research Center ‘Digital biodesign and personalized healthcare’ № 075-15-2022-304.

### Authors’ contributions

MB is the writer, researcher, collected and analyzed data, and revised the manuscript, EV collected the primary data from the hospital. All authors have read and approved the manuscript.

## Supporting information

Graphical abstract

## Data Availability

All data produced in the present study are available upon reasonable request to the authors

## Acknowledgments

not applicable

## Authors’ information

**Basheer Abdullah Marzoog**, Research Center «Digital Biodesign and Personalized Healthcare», I.M. Sechenov First Moscow State Medical University (Sechenov University), 119991 Moscow, Russia; postal address: Russia, Moscow, 8-2 Trubetskaya street, 119991. (, +79969602820). ORCID: 0000-0001-5507-2413. Scopus ID: 57486338800. **Ekaterina Vanichkina,** National Research Ogarev Mordovia State University. ORCID: 0009-0006-3015-2306

## The paper has not been submitted elsewhere

## Table Legend

Table 1: T-test results for grouping the sample by different principles to evaluate the potential role of each. Only statically significant parameters are shown in the table.

Table 2: Correlation between the numerical parameters of the sample. *Bold and italic values are statically significant correlations.

## Supplementary files

Supplementary file 1

The frequency table demonstrates the categorical variables in absolute and relative values.

Supplementary file 2

Graphical presentation of the correlation between the numerical parameters of the sample. All the graphs are statistically significant.

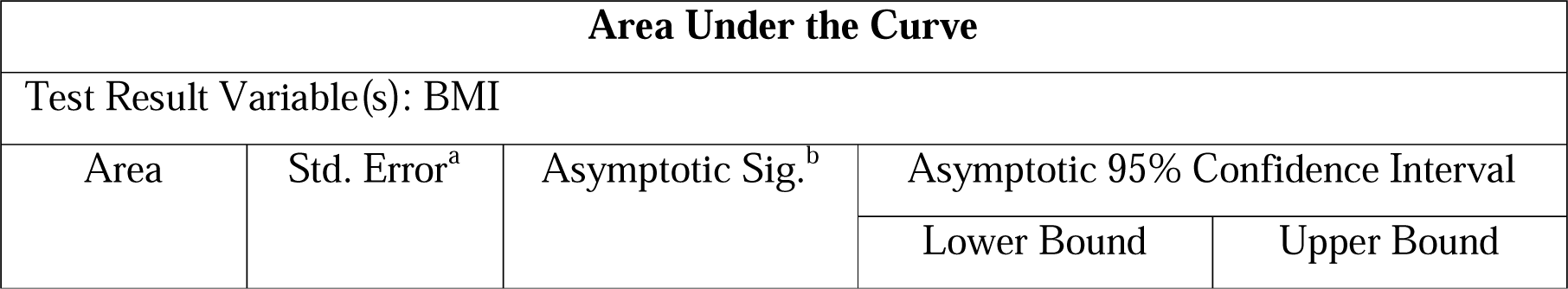
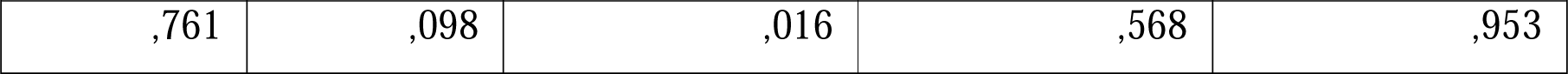

ROC analyses for the cut-point body mass index (BMI) in males as a gender risk factor for myocardial infarction. a. Under the nonparametric assumption. b. Null hypothesis: true area = 0.5

**Figure 1:**
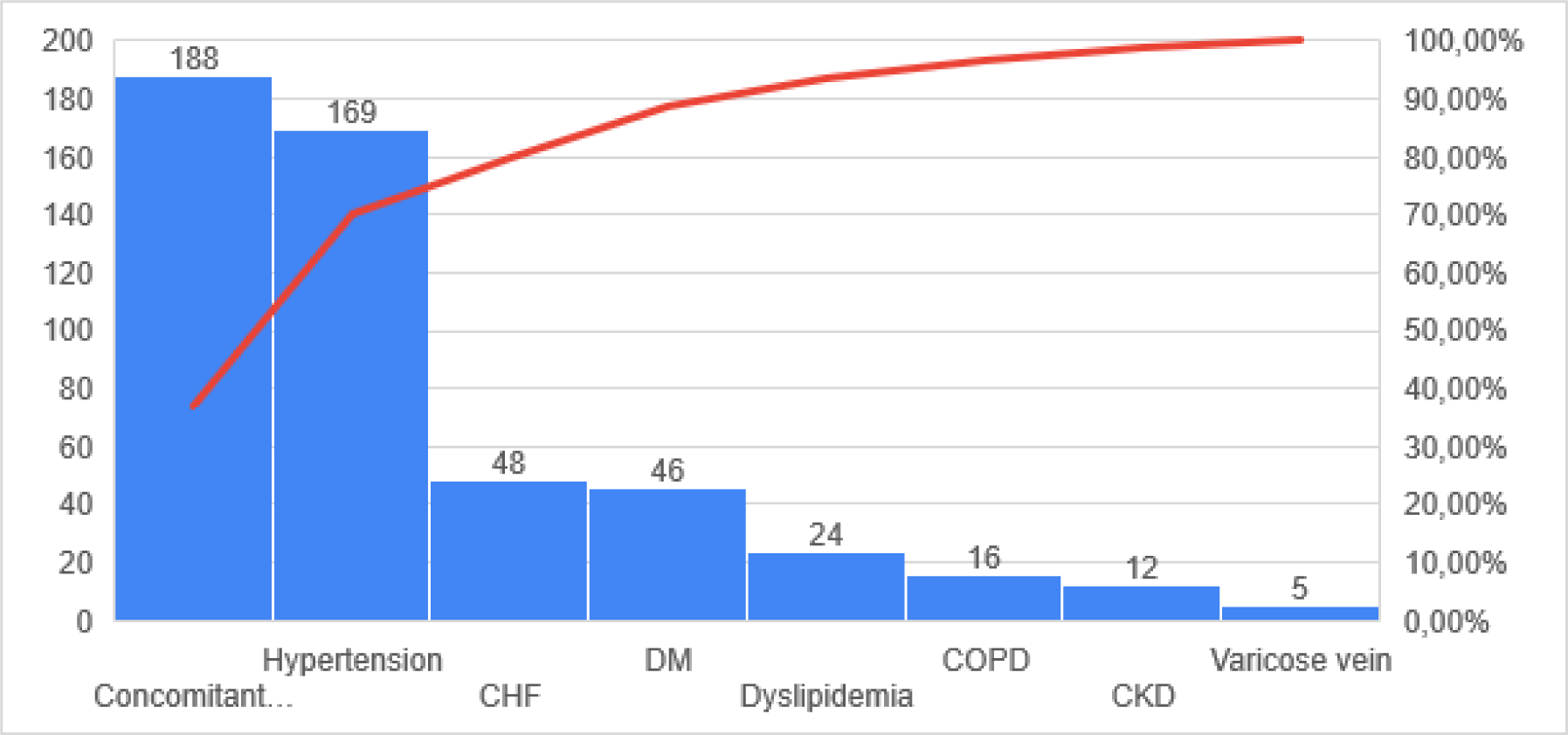
Most frequently concomitant diseases reported in MI victims. descending arrange for the concomitant diseases by frequency of occurrence. Abbreviations: CHF; chronic heart failure, DM; diabetes mellitus, COPD; chronic obstructive lung disease, CKD; chronic kidney disease.

**Figure 1:**
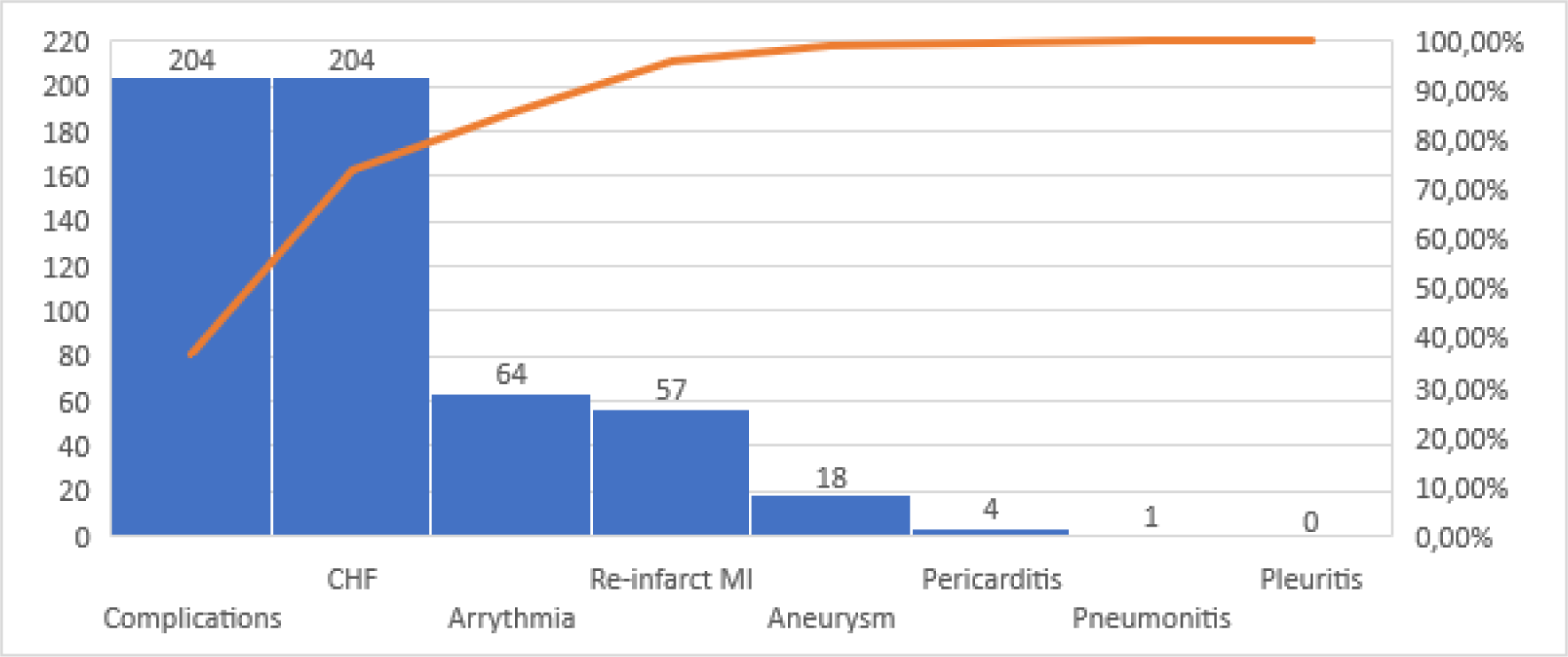
Most frequently reported complications post PCI and MI in MI victims. Descending arranges for the complications by frequency of occurrence. Abbreviations CHF; chronic heart failure, MI; myocardial infarction.

**Figure 1:**
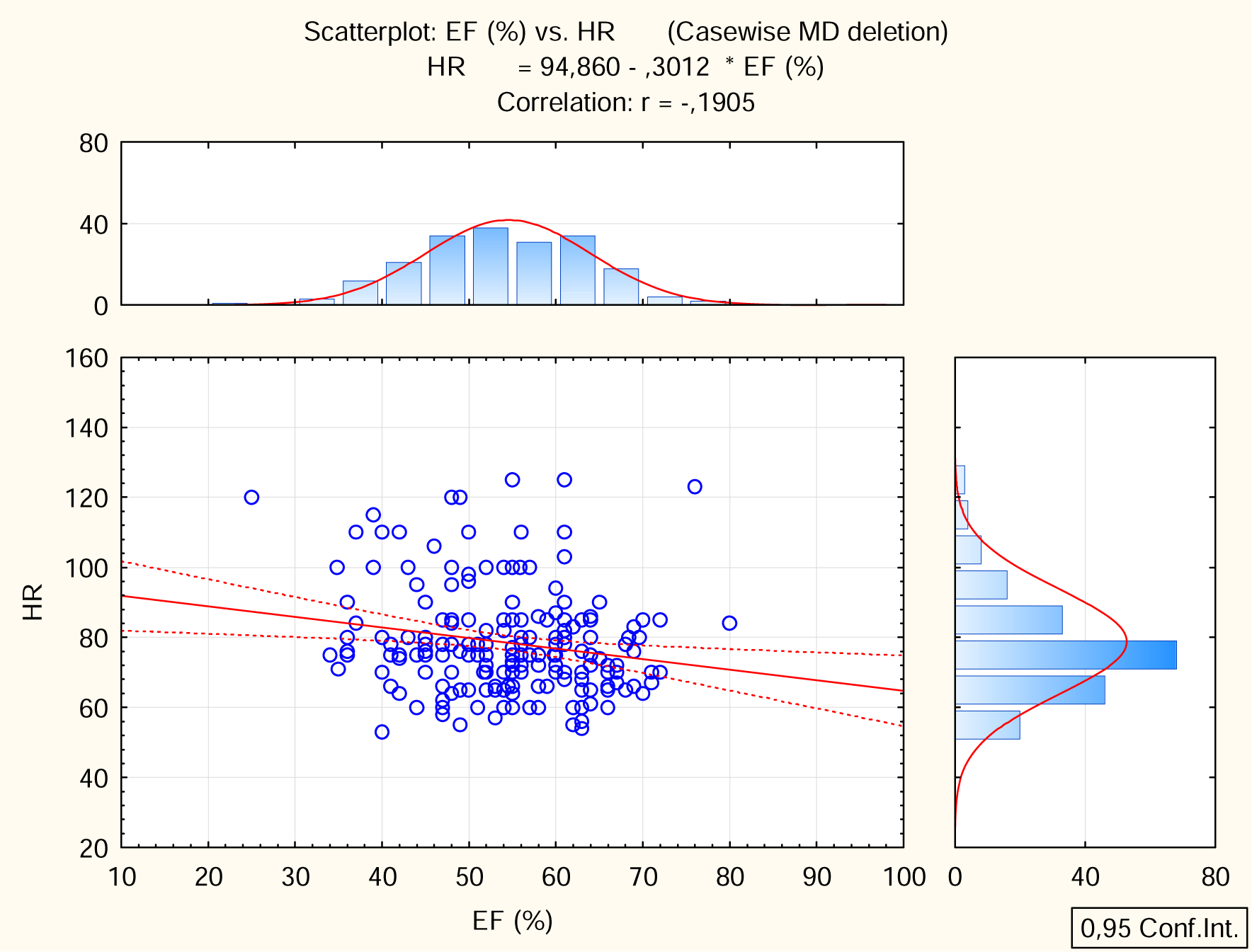
Proportional correlation between the heart rate and the ejection fraction.

**Figure 1:**
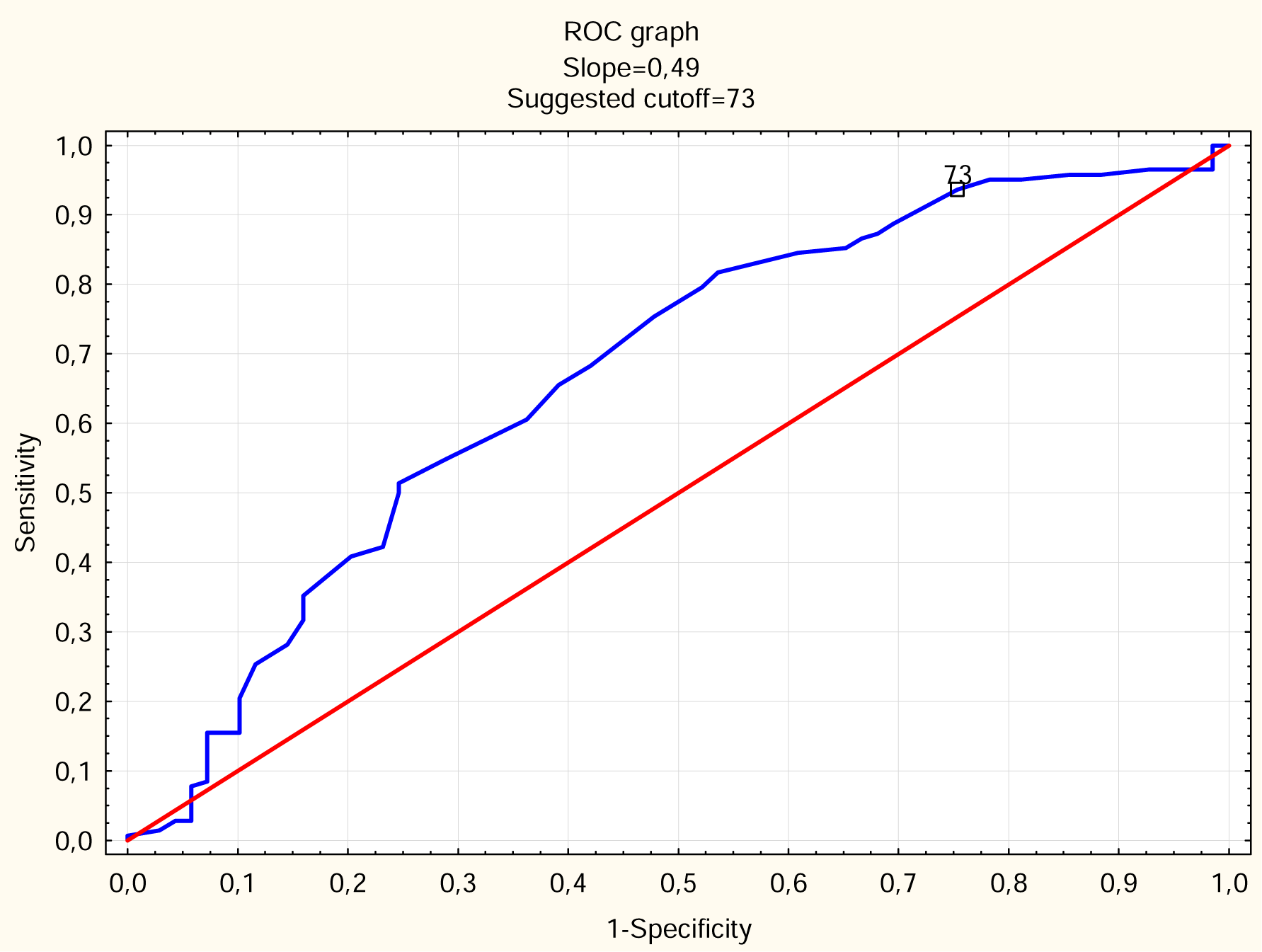
ROC analyses for the cut-point age in females as a gender risk factor.

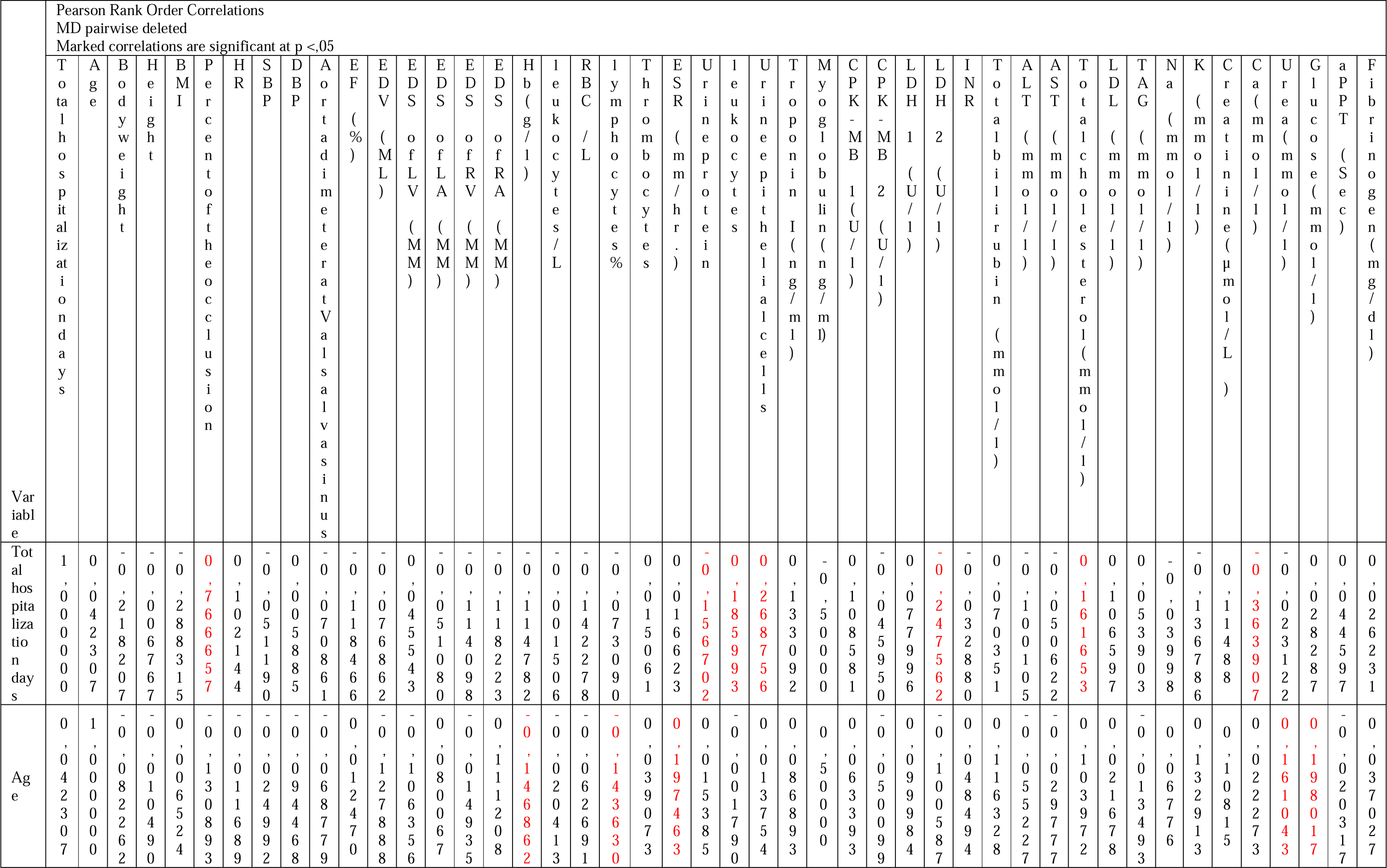
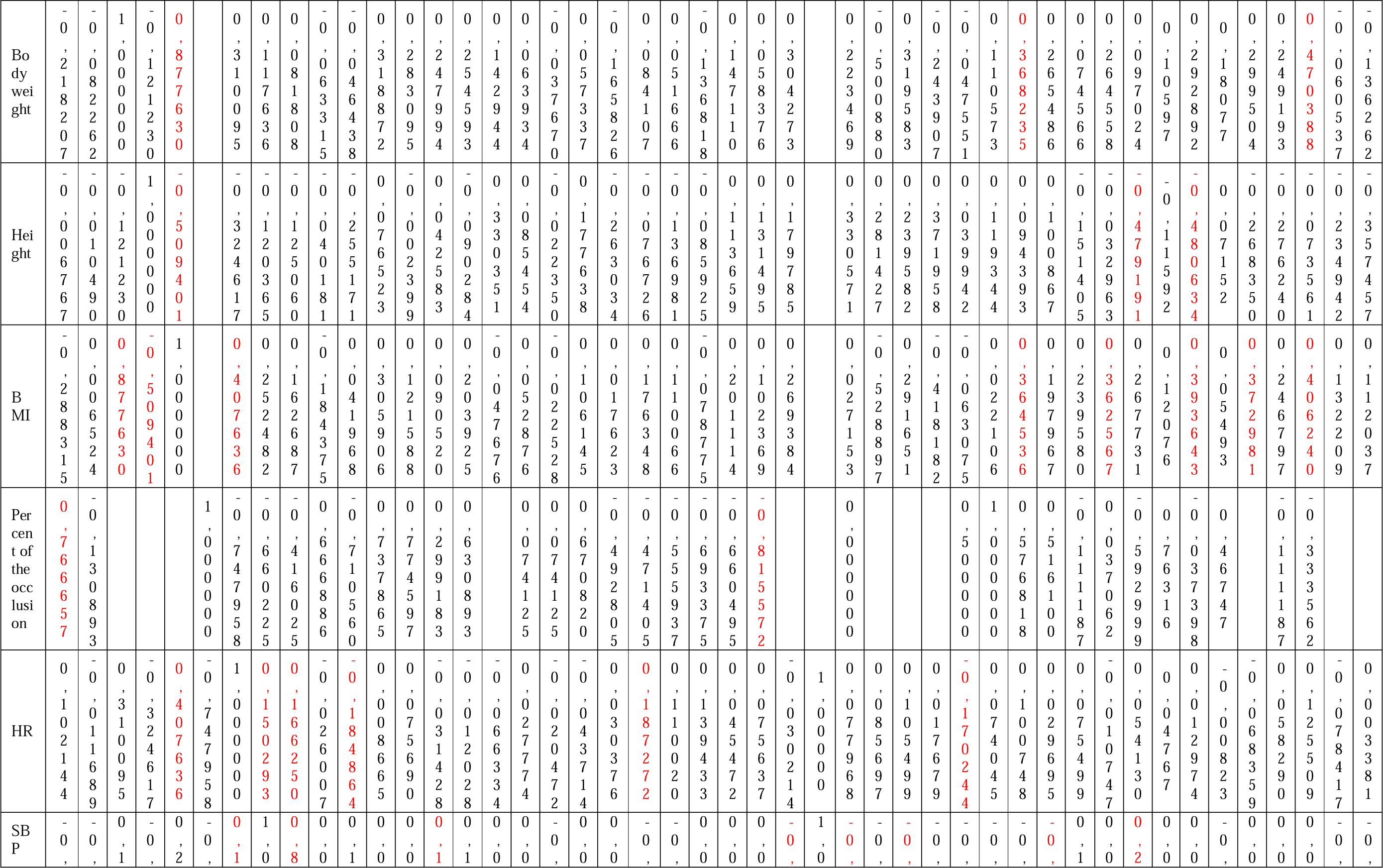
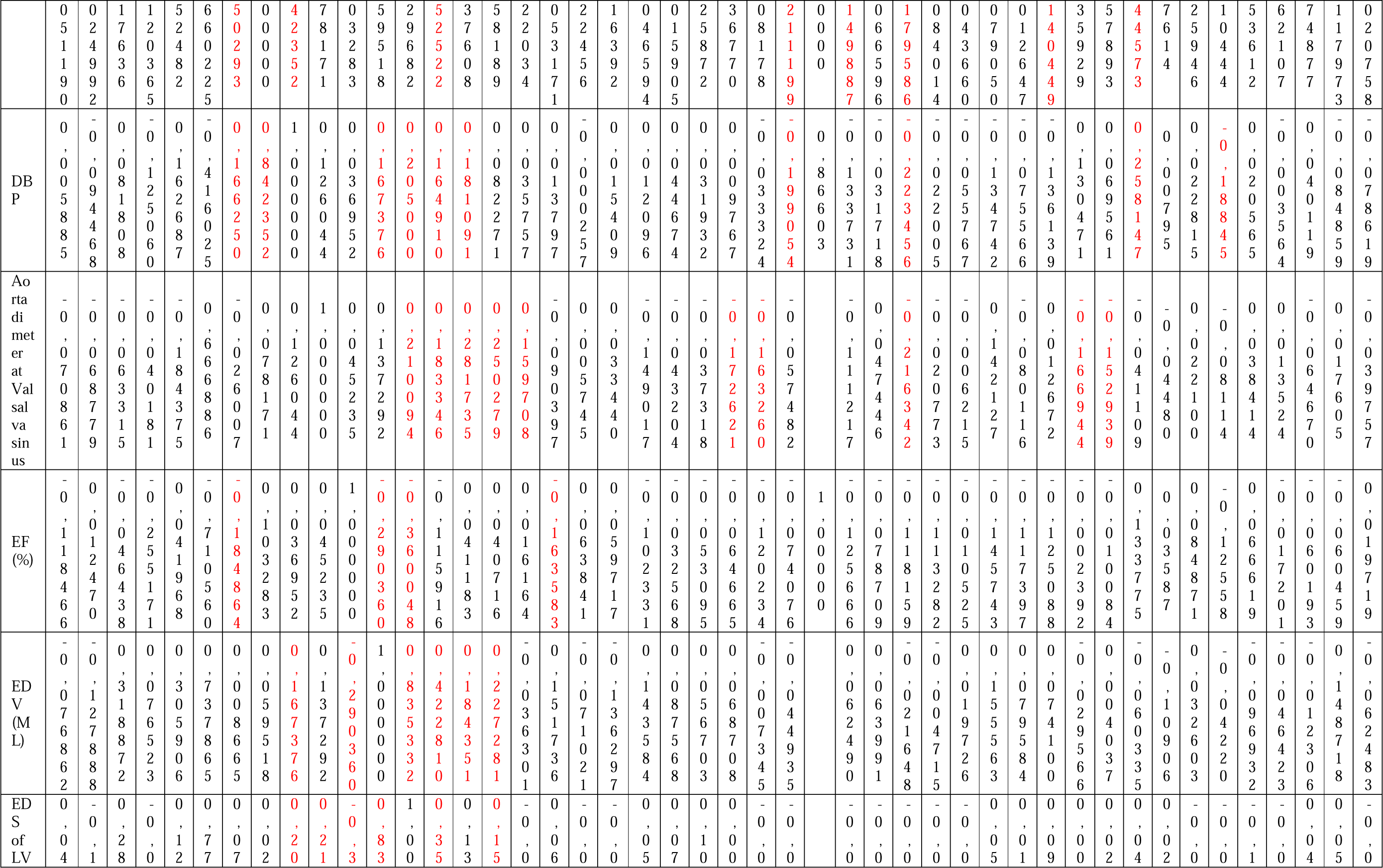
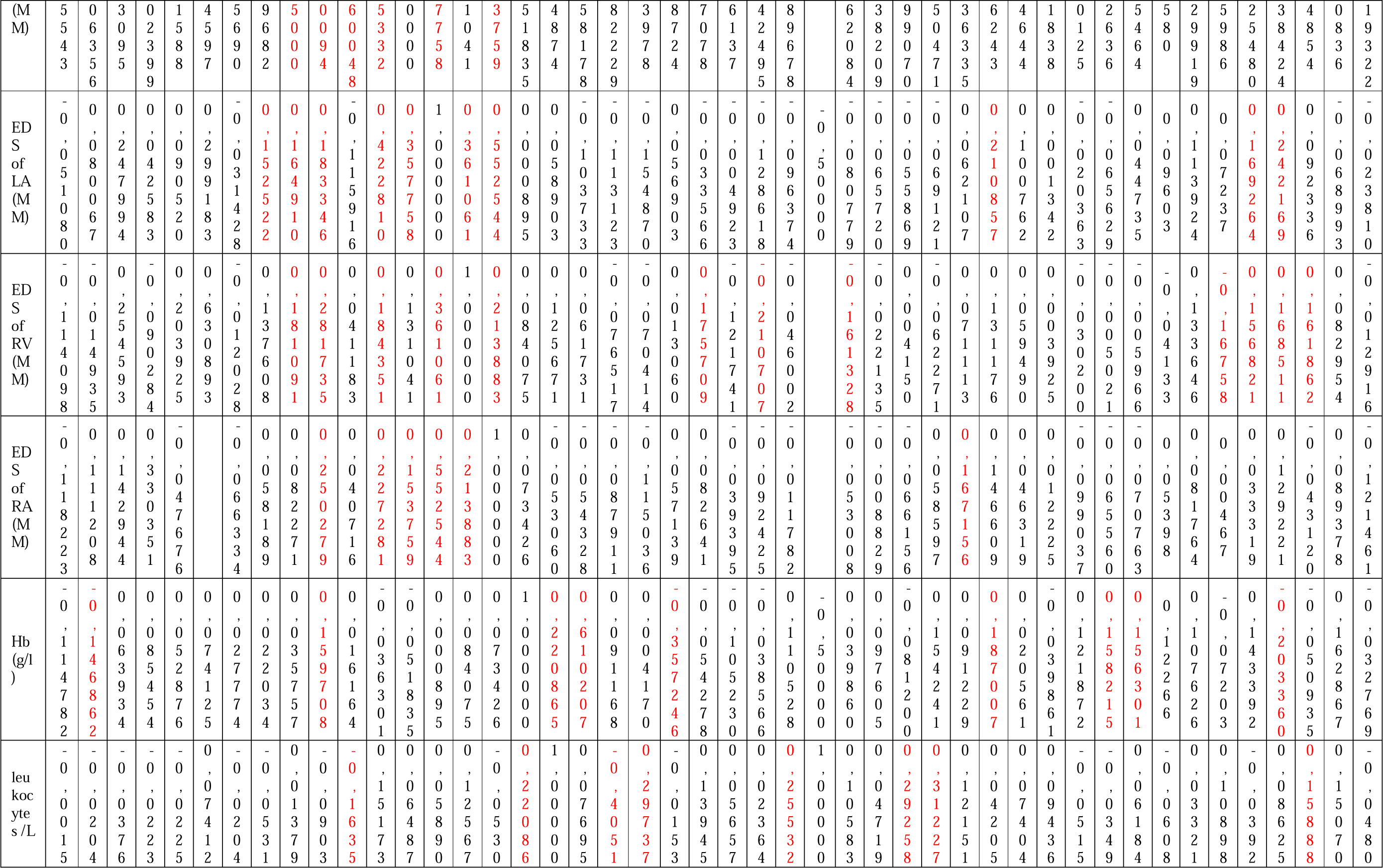
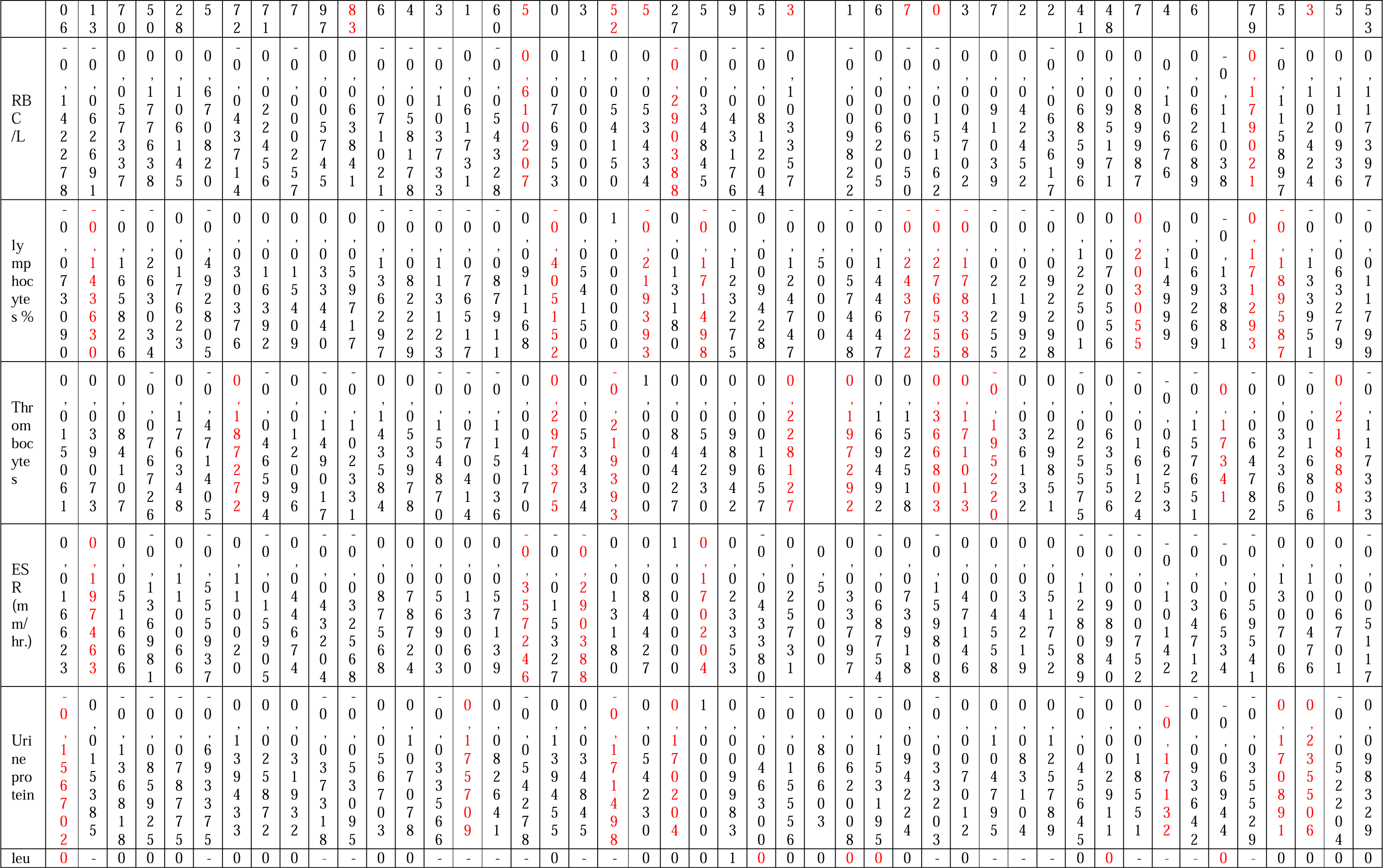
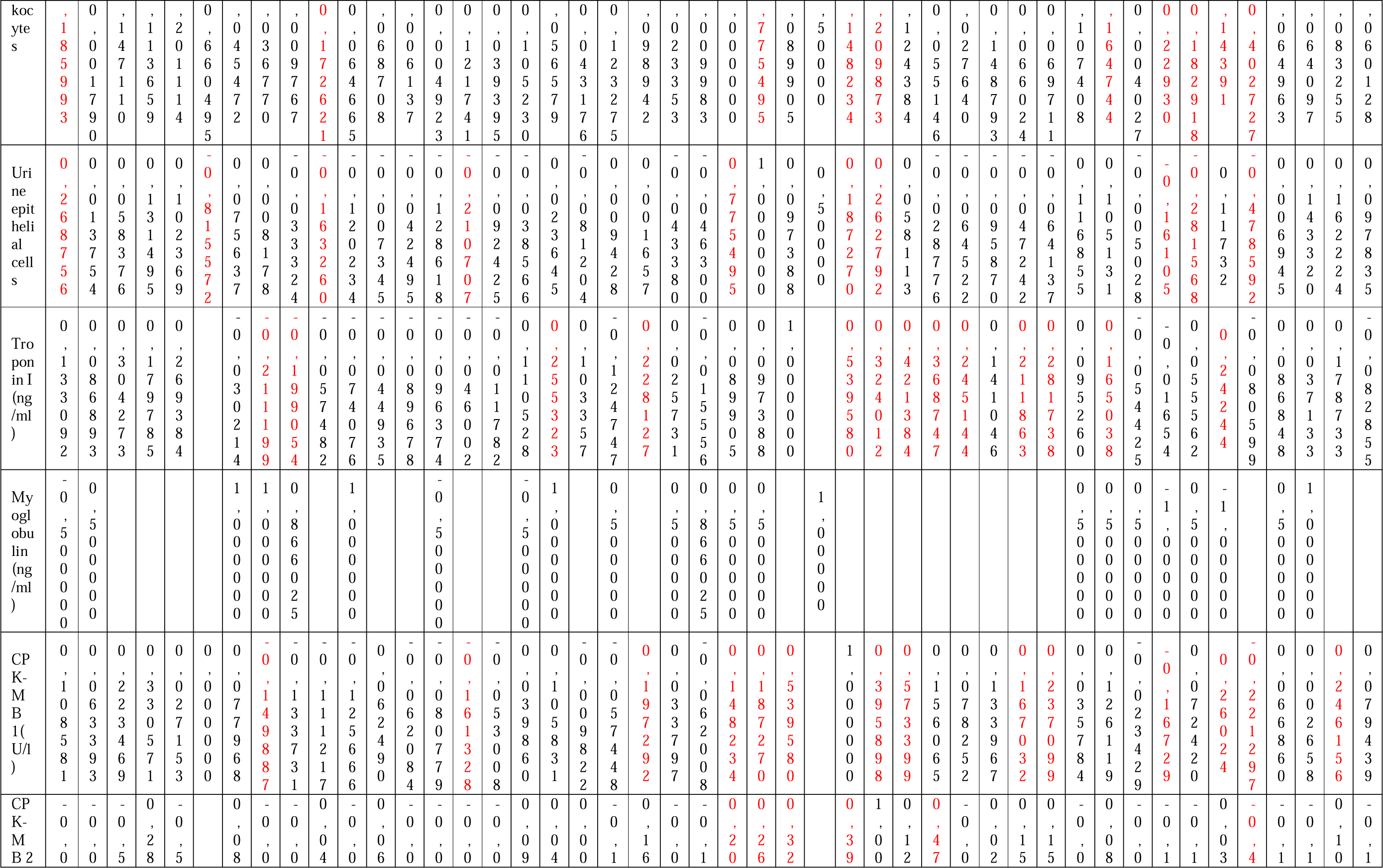
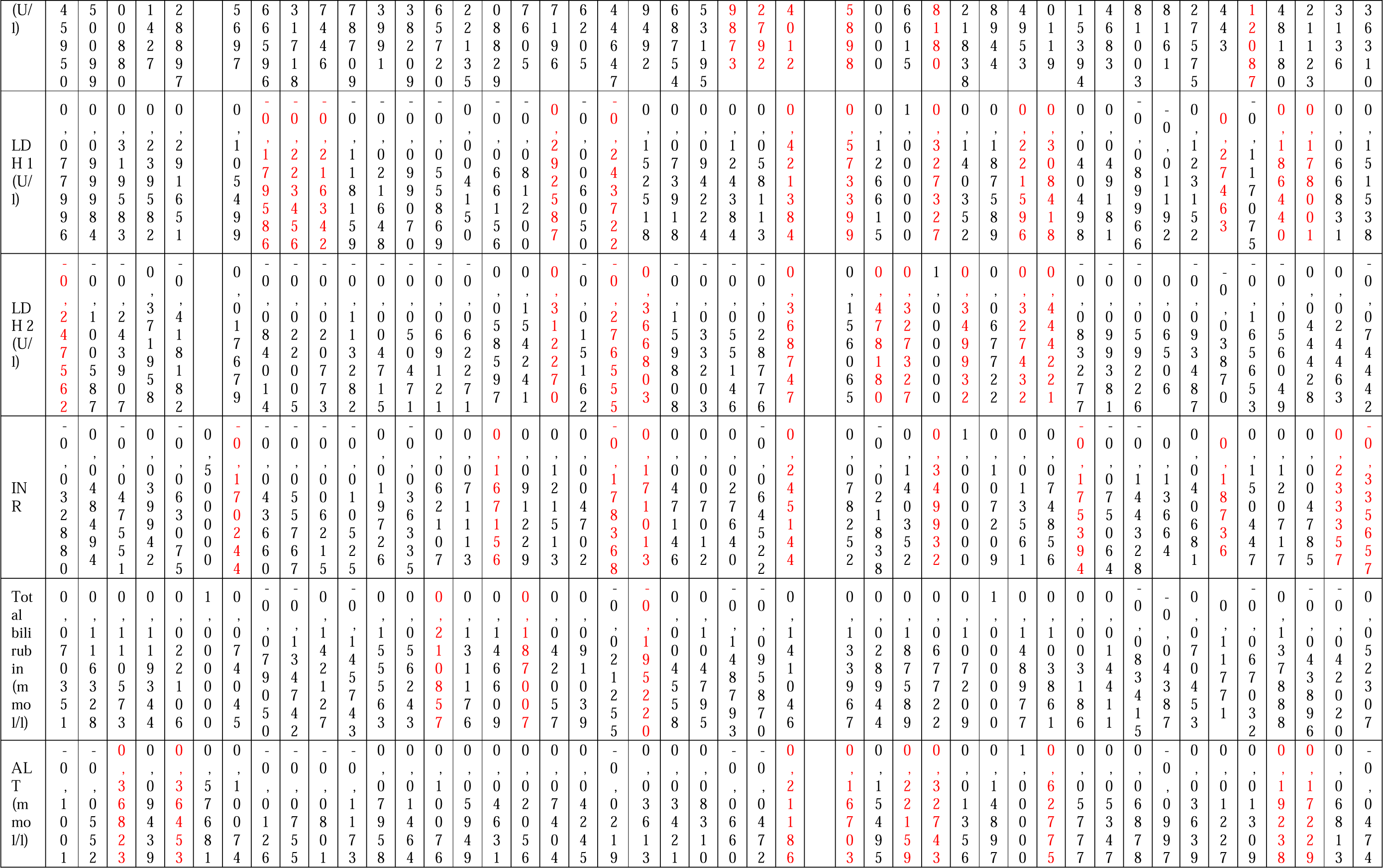
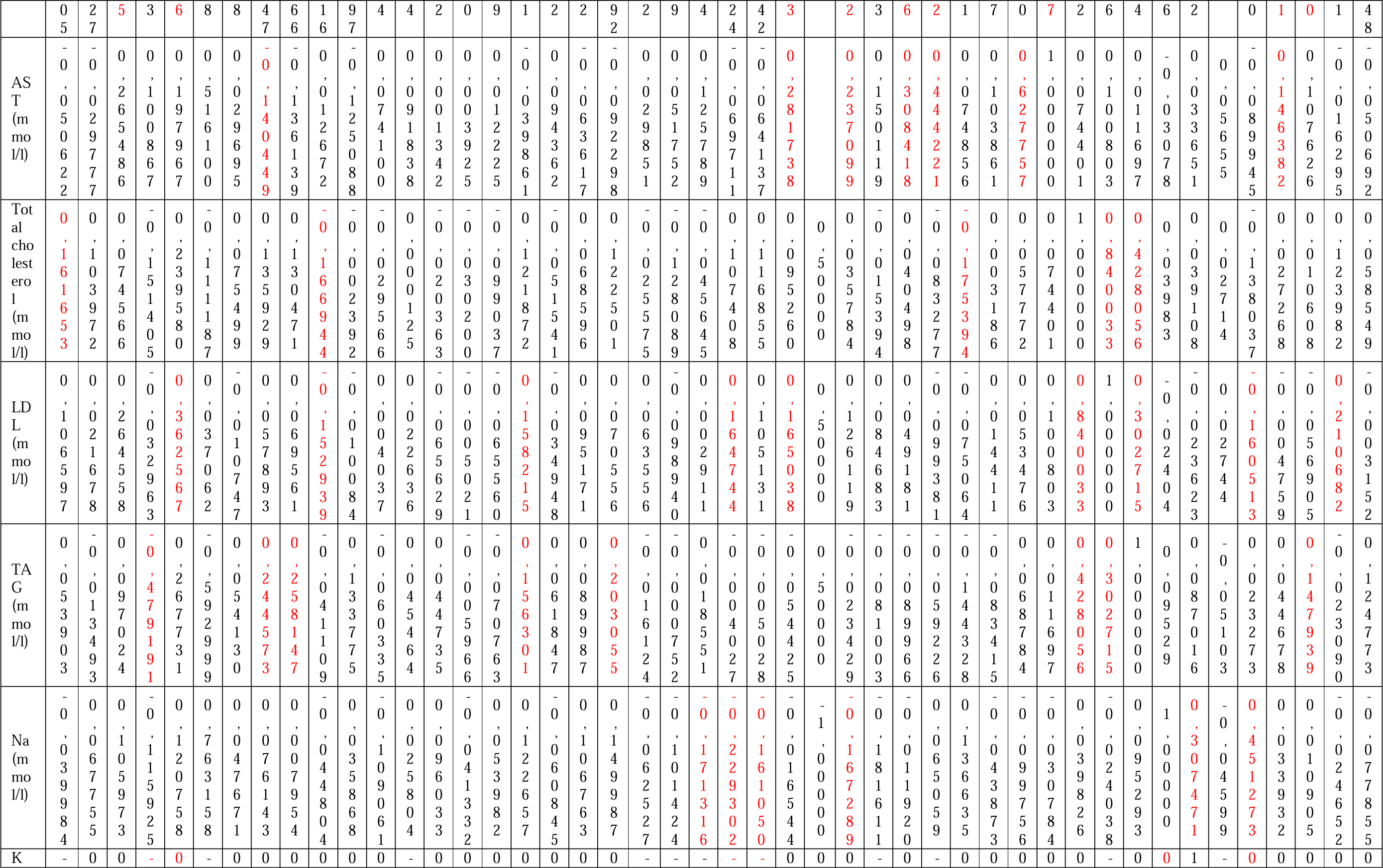
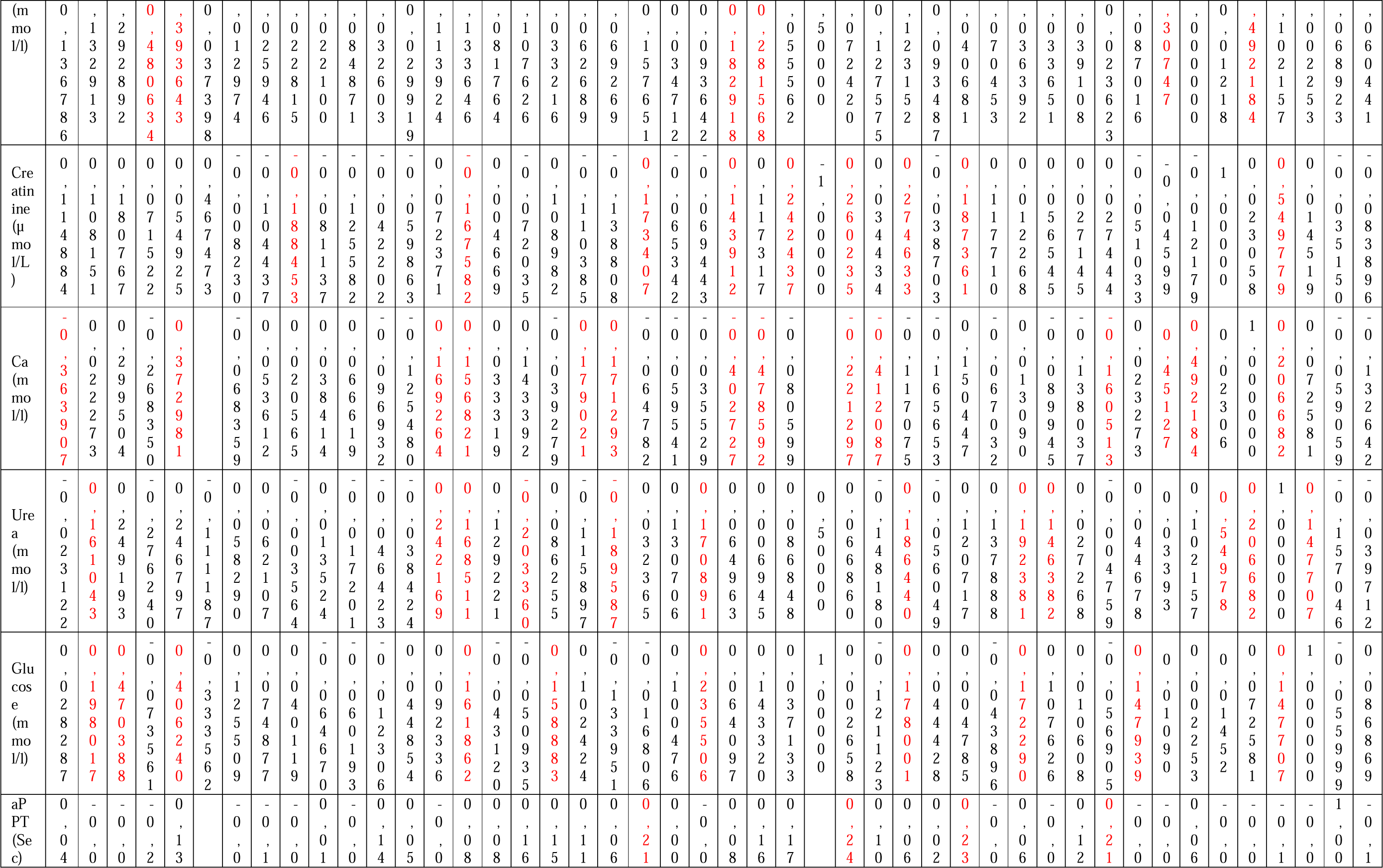
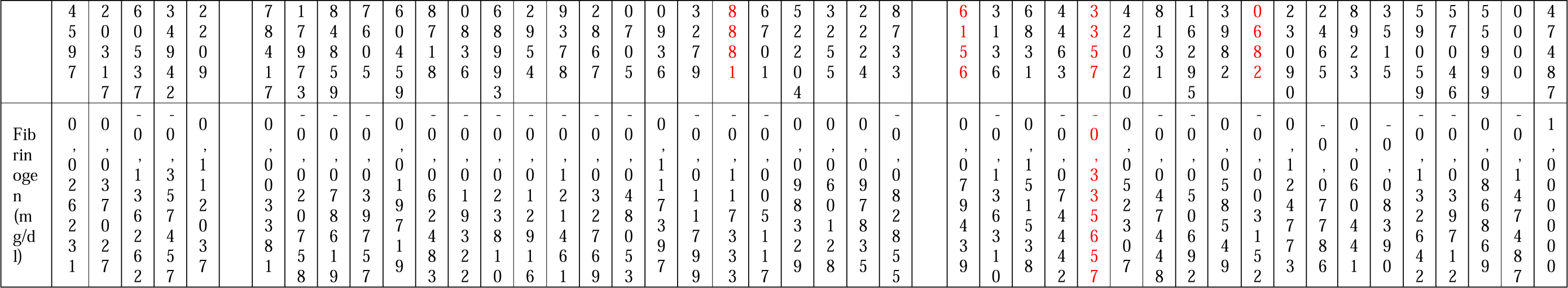

## Notes

### Competing Interest Statement

The authors have declared no competing interest.

### Author Declarations

The study approved by the National Research Mordovia State University, Russia, from Ethics Committee Requirement N8/2 from 30.06.2022.

### Summary of Updates

Updated the tittle of the paper, updated the author list to include " Ekaterina Vanichkina ". also some changes carred out on the text and grammaticla corrections.

