## Supplementary material for "Early Prognostic Instrumental and Laboratory Biomarkers in Post-MI": Graphical abstract

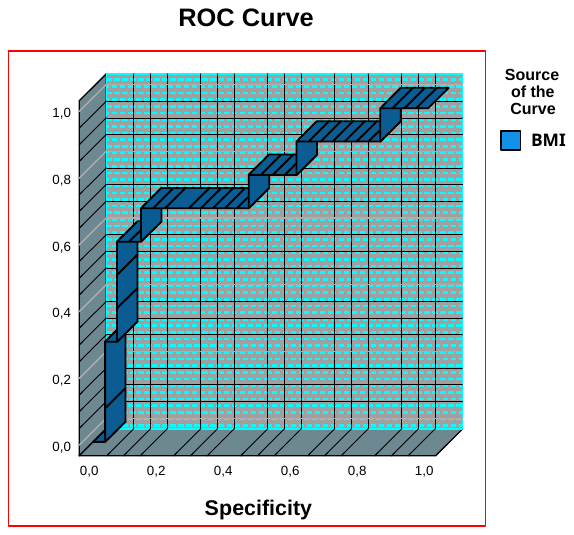

| **Area Under the Curve** | | | | |
| --- | --- | --- | --- | --- |
| Test Result Variable(s): BMI | | | | |
| Area | Std. Error^a^ | Asymptotic Sig.^b^ | Asymptotic 95% Confidence Interval | |
|  |  |  | Lower Bound | Upper Bound |
| ,761 | ,098 | ,016 | ,568 | ,953 |

ROC analyses for the cut-point body mass index (BMI) in males as a gender risk factor for myocardial infarction. a. Under the nonparametric assumption. b. Null hypothesis: true area = 0.5
