## Supplementary material for "Early Prognostic Instrumental and Laboratory Biomarkers in Post-MI": Supp. 1

| Category | Frequency table: Gender (Acute MI in Total Post MI final. stw) | | | |
| --- | --- | --- | --- | --- |
|  | Count | Cumulative  Count | Percent | Cumulative  Percent |
| F | 69 | 69 | 32,70142 | 32,7014 |
| M | 142 | 211 | 67,29858 | 100,0000 |
| Missing | 0 | 211 | 0,00000 | 100,0000 |

Gender distribution in the study.

The frequency of patients with concomitant disease.

| Category | Frequency table: Concomitant disease (Acute MI in Total Post MI final. stw) | | | |
| --- | --- | --- | --- | --- |
|  | Count | Cumulative  Count | Percent | Cumulative  Percent |
| Yes | 188 | 188 | 89,09953 | 89,0995 |
| No | 22 | 210 | 10,42654 | 99,5261 |
| Missing | 1 | 211 | 0,47393 | 100,0000 |

| Category | Frequency table: CHF (Acute MI in Total Post MI final. stw) | | | |
| --- | --- | --- | --- | --- |
|  | Count | Cumulative  Count | Percent | Cumulative  Percent |
| No | 154 | 154 | 72,98578 | 72,9858 |
| Yes | 49 | 203 | 23,2227 | 96,2085 |
| Missing | 8 | 211 | 3,79147 | 100,0000 |

| Category | Frequency table: DM (Acute MI in Total Post MI final. stw) | | | |
| --- | --- | --- | --- | --- |
|  | Count | Cumulative  Count | Percent | Cumulative  Percent |
| No | 160 | 160 | 75,82938 | 75,8294 |
| Yes | 46 | 206 | 21,80095 | 97,6303 |
| Missing | 5 | 211 | 2,36967 | 100,0000 |

| Category | Frequency table: Hypertension (Acute MI in Total Post MI final. stw) | | | |
| --- | --- | --- | --- | --- |
|  | Count | Cumulative  Count | Percent | Cumulative  Percent |
| Yes | 169 | 169 | 80,09479 | 80,0948 |
| No | 37 | 206 | 17,53555 | 97,6303 |
| Missing | 5 | 211 | 2,36967 | 100,0000 |

| Category | Frequency table: CKD (Acute MI in Total Post MI final. stw) | | | |
| --- | --- | --- | --- | --- |
|  | Count | Cumulative  Count | Percent | Cumulative  Percent |
| No | 189 | 189 | 89,57346 | 89,5735 |
| Yes | 12 | 201 | 6,1611 | 95,7346374 |
| Missing | 9 | 211 | 4,26540 | 100,0000 |

| Category | Frequency table: COPD (Acute MI in Total Post MI final. stw) | | | |
| --- | --- | --- | --- | --- |
|  | Count | Cumulative  Count | Percent | Cumulative  Percent |
| No | 190 | 190 | 90,04739 | 90,0474 |
| Yes | 16 | 206 | 7,58294 | 97,6303 |
| Missing | 5 | 211 | 2,36967 | 100,0000 |

| Category | Frequency table: Dyslipidemia (Acute MI in Total Post MI final. stw) | | | |
| --- | --- | --- | --- | --- |
|  | Count | Cumulative  Count | Percent | Cumulative  Percent |
| No | 181 | 181 | 85,78199 | 85,7820 |
| Yes | 24 | 205 | 11,37441 | 97,1564 |
| Missing | 6 | 211 | 2,84360 | 100,0000 |

| Category | Frequency table: Varicose vein (Acute MI in Total Post MI final. stw) | | | |
| --- | --- | --- | --- | --- |
|  | Count | Cumulative  Count | Percent | Cumulative  Percent |
| No | 200 | 200 | 94,78673 | 94,7867 |
| Yes | 5 | 205 | 2,36967 | 97,1564 |
| Missing | 6 | 211 | 2,84360 | 100,0000 |

The frequency of post myocardial infarction complications.

| Category | Frequency table: Complications (Acute MI in Total Post MI final. stw) | | | |
| --- | --- | --- | --- | --- |
|  | Count | Cumulative  Count | Percent | Cumulative  Percent |
| Yes | 202 | 202 | 95,73460 | 95,7346 |
| No | 7 | 209 | 3,31754 | 99,0521 |
| Missing | 2 | 211 | 0,94787 | 100,0000 |

| Category | Frequency table: CHF (Acute MI in Total Post MI final. stw) | | | |
| --- | --- | --- | --- | --- |
|  | Count | Cumulative  Count | Percent | Cumulative  Percent |
| Yes | 204 | 204 | 96,68246 | 96,6825 |
| No | 4 | 208 | 1,89573 | 98,5782 |
| Missing | 3 | 211 | 1,42180 | 100,0000 |

| Category | Frequency table: Arrythmia (Acute MI in Total Post MI final. stw) | | | |
| --- | --- | --- | --- | --- |
|  | Count | Cumulative  Count | Percent | Cumulative  Percent |
| Yes | 66 | 66 | 31,2796 | 31,2796 |
| No | 143 | 209 | 67,77251 | 99,0521309 |
| Missing | 2 | 211 | 0,94787 | 100,0000 |

| Category | Frequency table: Re-infarct MI (Acute MI in Total Post MI final. stw) | | | |
| --- | --- | --- | --- | --- |
|  | Count | Cumulative  Count | Percent | Cumulative  Percent |
| Yes | 57 | 57 | 27,01422 | 27,0142 |
| No | 153 | 210 | 72,51185 | 99,5261 |
| Missing | 1 | 211 | 0,47393 | 100,0000 |

| Category | Frequency table: Aneurysm (Acute MI in Total Post MI final. stw) | | | |
| --- | --- | --- | --- | --- |
|  | Count | Cumulative  Count | Percent | Cumulative  Percent |
| No | 192 | 192 | 90,99526 | 90,9953 |
| Yes | 18 | 210 | 8,53081 | 99,5261 |
| Missing | 1 | 211 | 0,47393 | 100,0000 |

| Category | Frequency table: Pericarditis (Acute MI in Total Post MI final. stw) | | | |
| --- | --- | --- | --- | --- |
|  | Count | Cumulative  Count | Percent | Cumulative  Percent |
| No | 206 | 206 | 97,63033 | 97,6303 |
| Yes | 4 | 210 | 1,89573 | 99,5261 |
| Missing | 1 | 211 | 0,47393 | 100,0000 |

| Category | Frequency table: Pneumonitis (Acute MI in Total Post MI final. stw) | | | |
| --- | --- | --- | --- | --- |
|  | Count | Cumulative  Count | Percent | Cumulative  Percent |
| No | 208 | 208 | 98,57820 | 98,5782 |
| Yes | 1 | 209 | 0,47393 | 99,0521 |
| Missing | 2 | 211 | 0,94787 | 100,0000 |

| Category | Frequency table: Pleuritis (Acute MI in Total Post MI final. stw) | | | |
| --- | --- | --- | --- | --- |
|  | Count | Cumulative  Count | Percent | Cumulative  Percent |
| No | 210 | 210 | 99,52607 | 99,5261 |
| Missing | 1 | 211 | 0,47393 | 100,0000 |
