## Supplementary material for "Early Prognostic Instrumental and Laboratory Biomarkers in Post-MI": Supp. 2

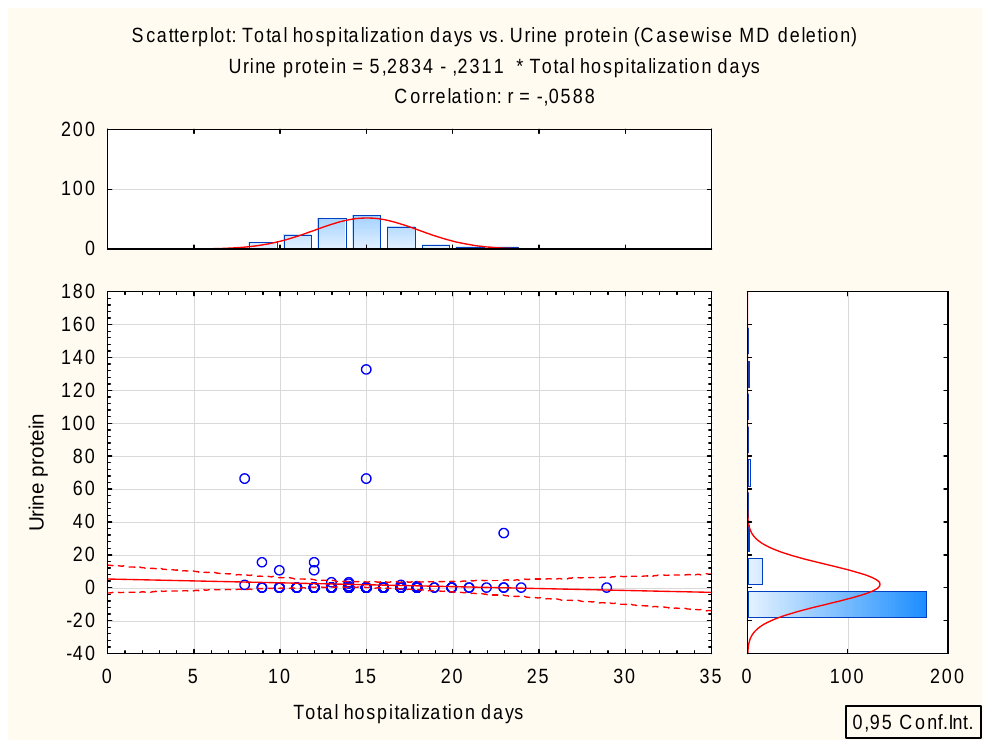

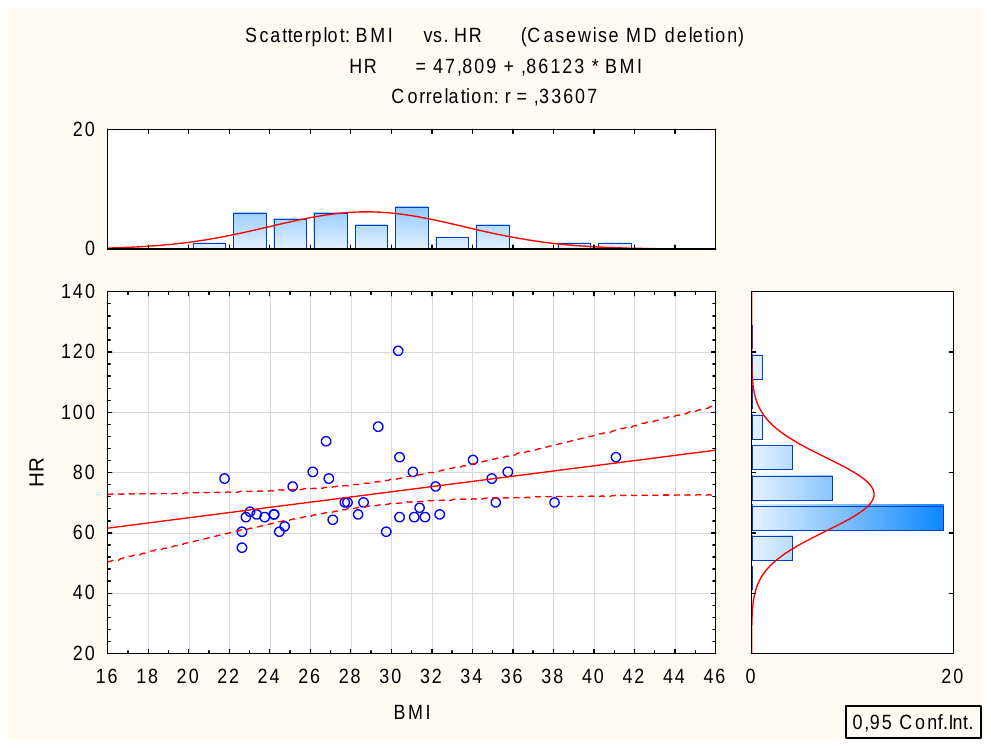

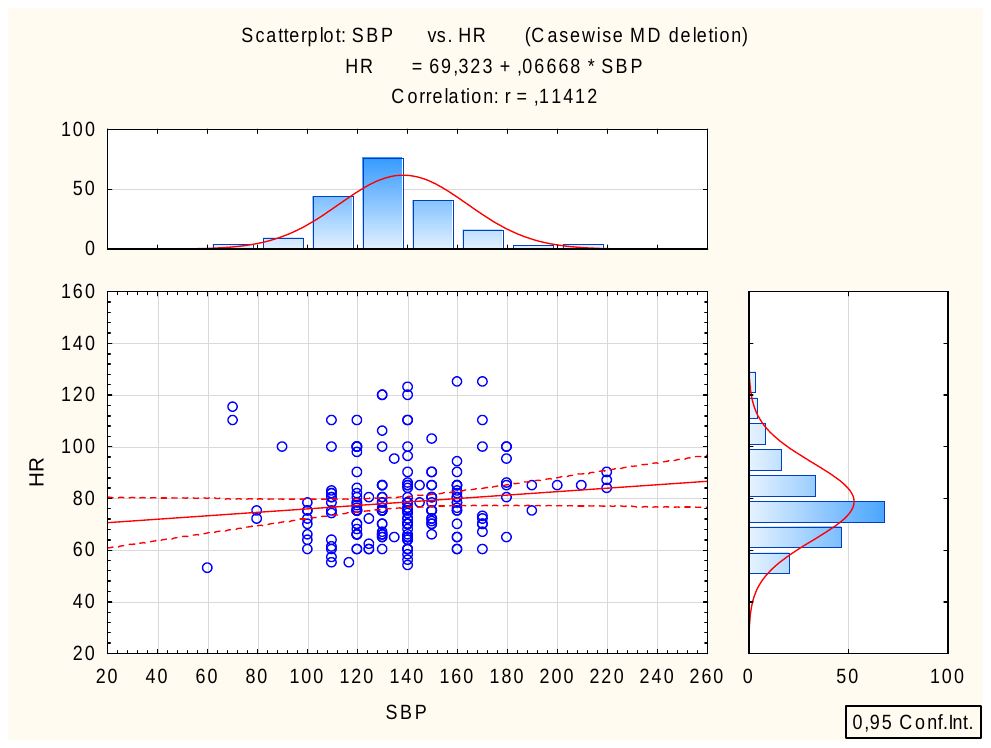

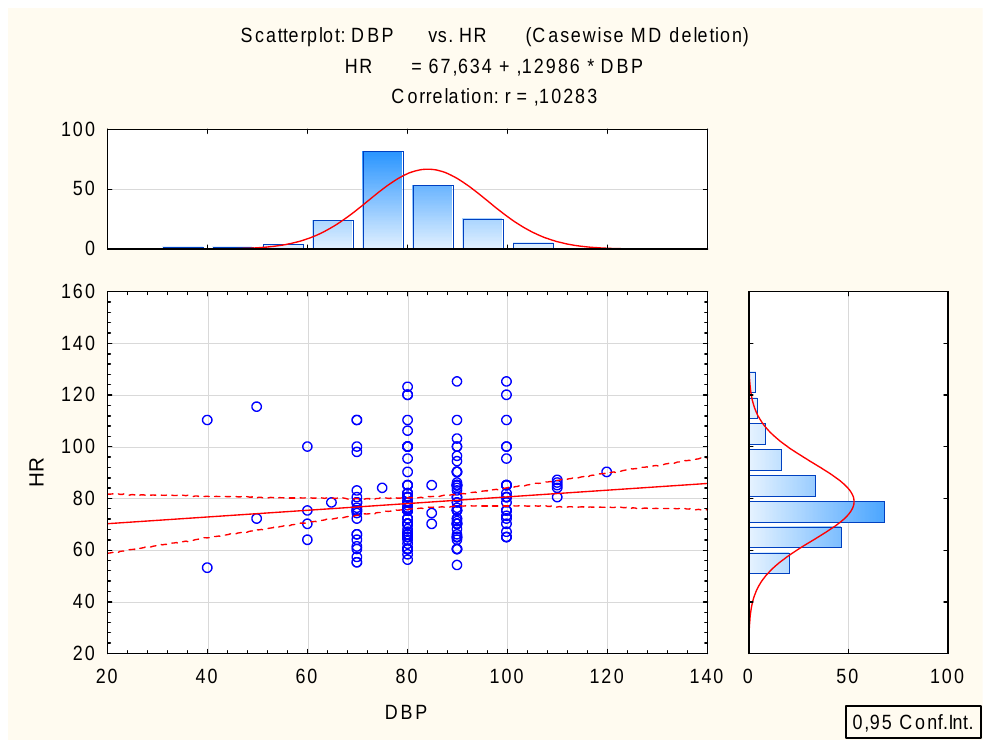

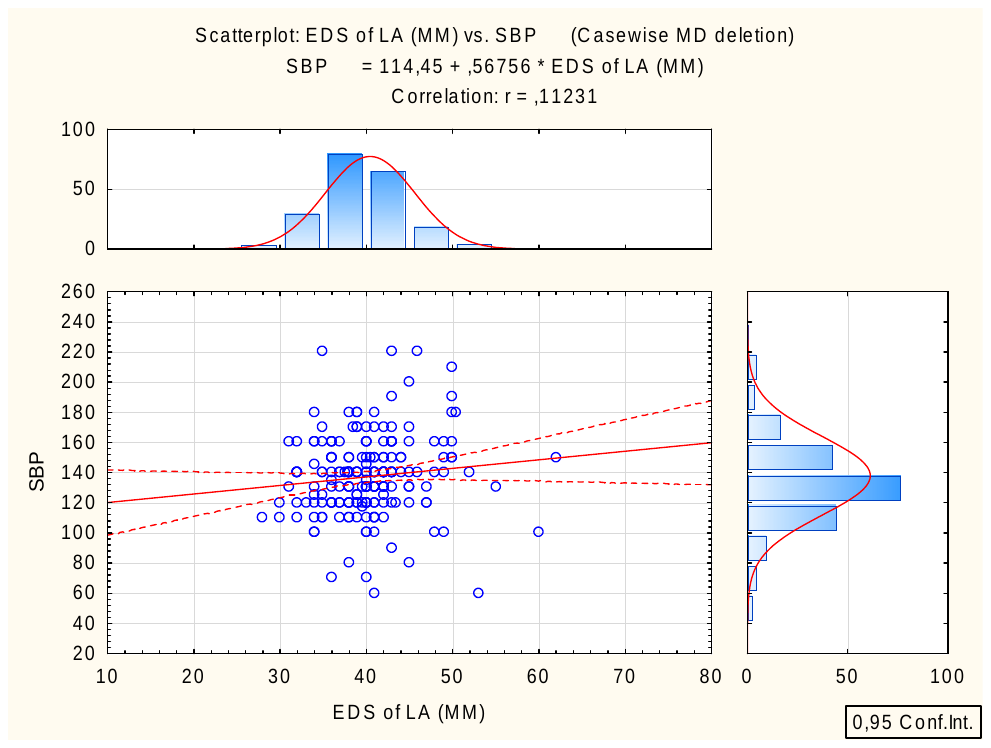

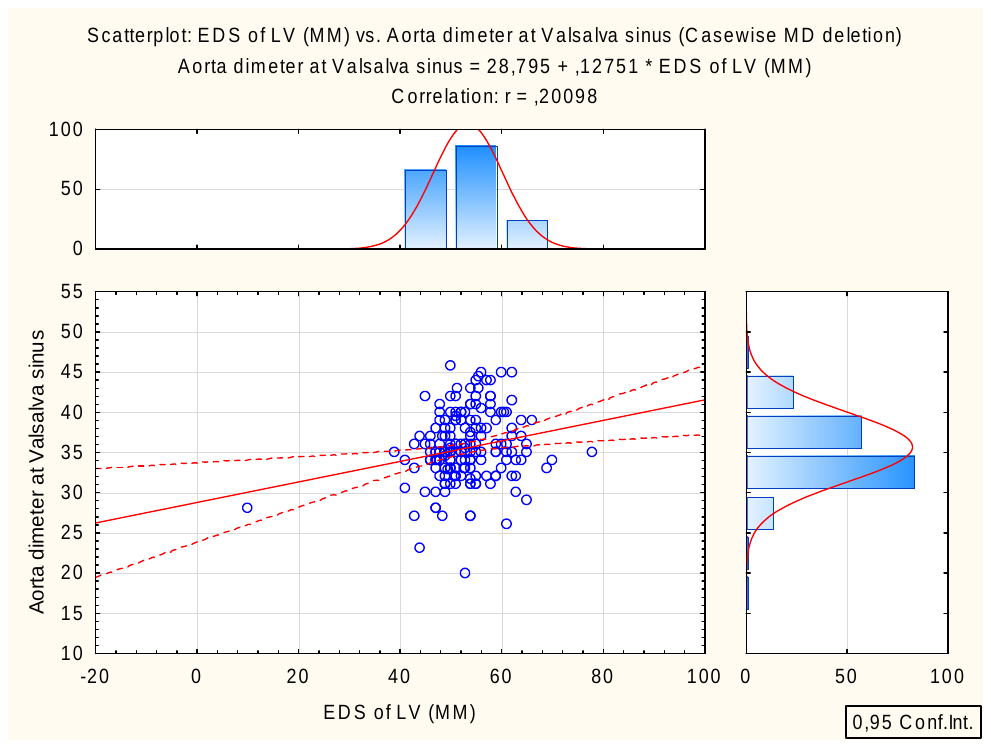

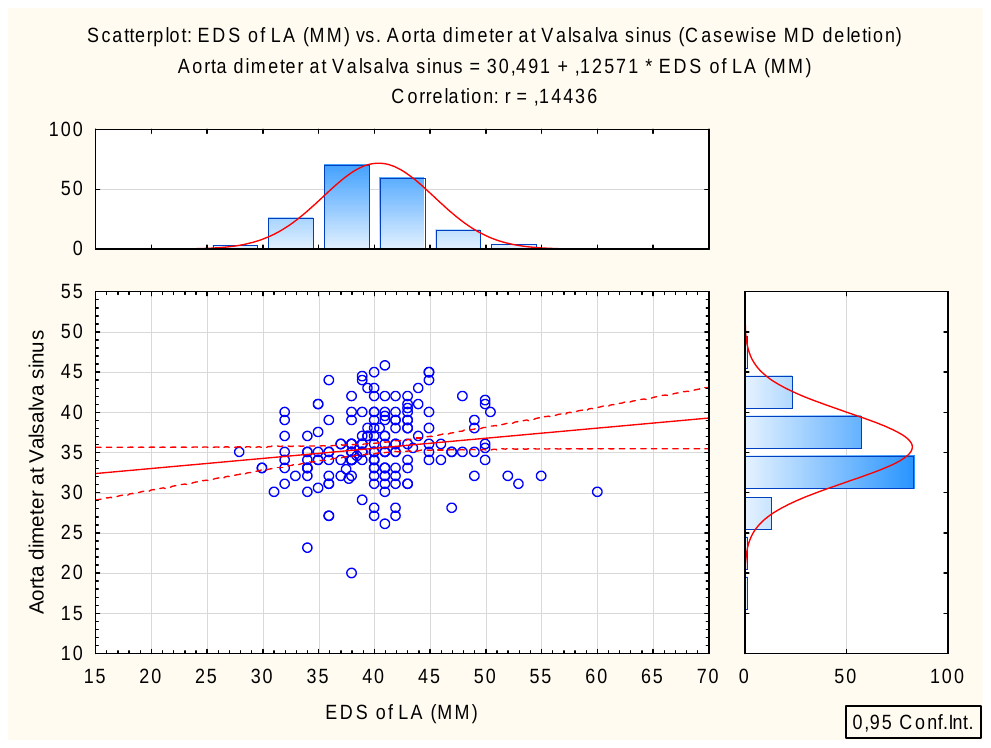

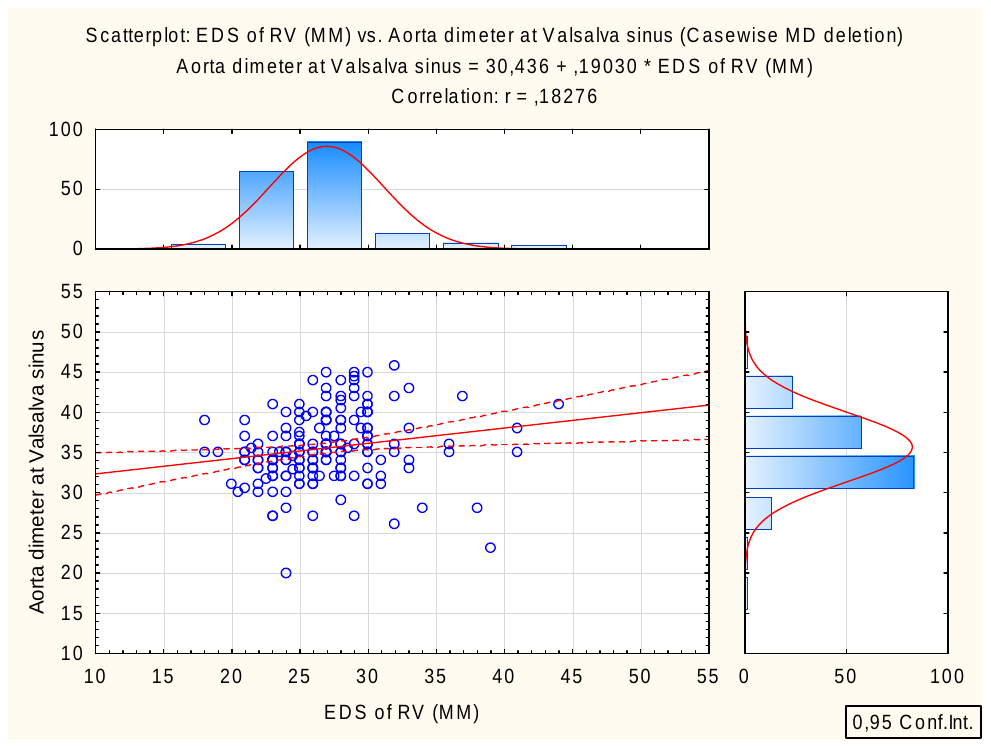

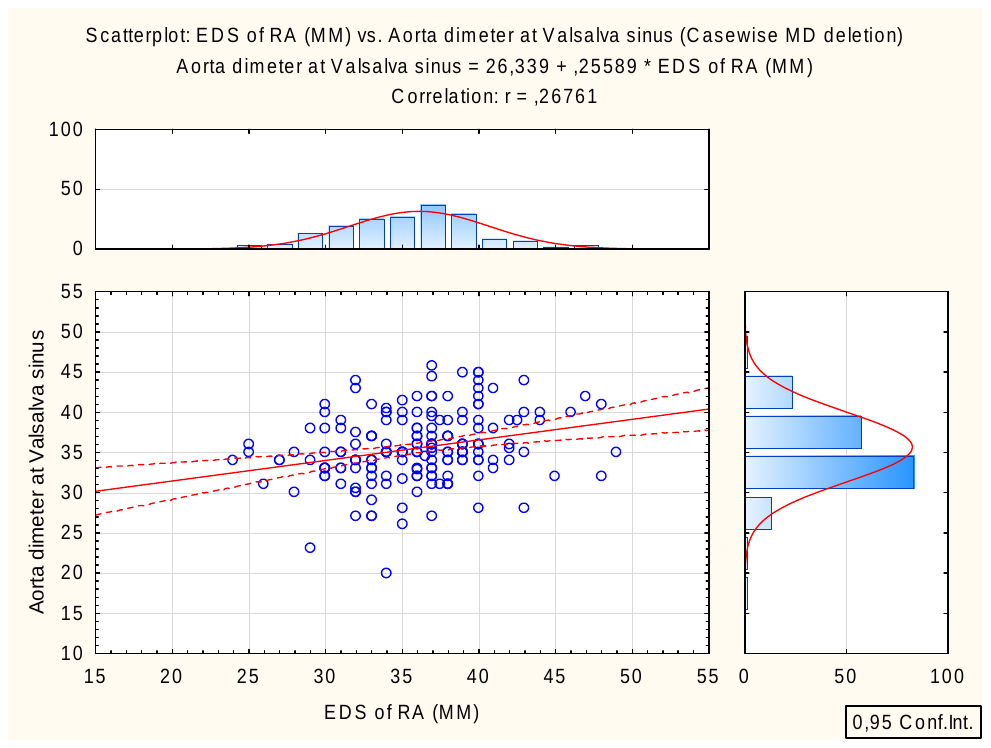

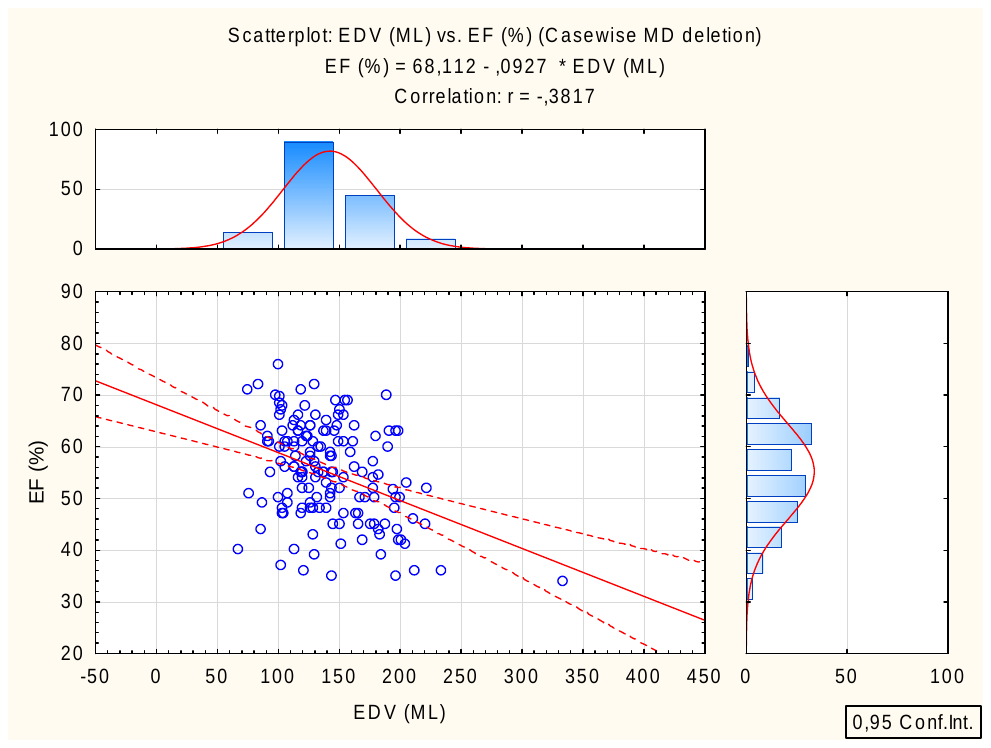

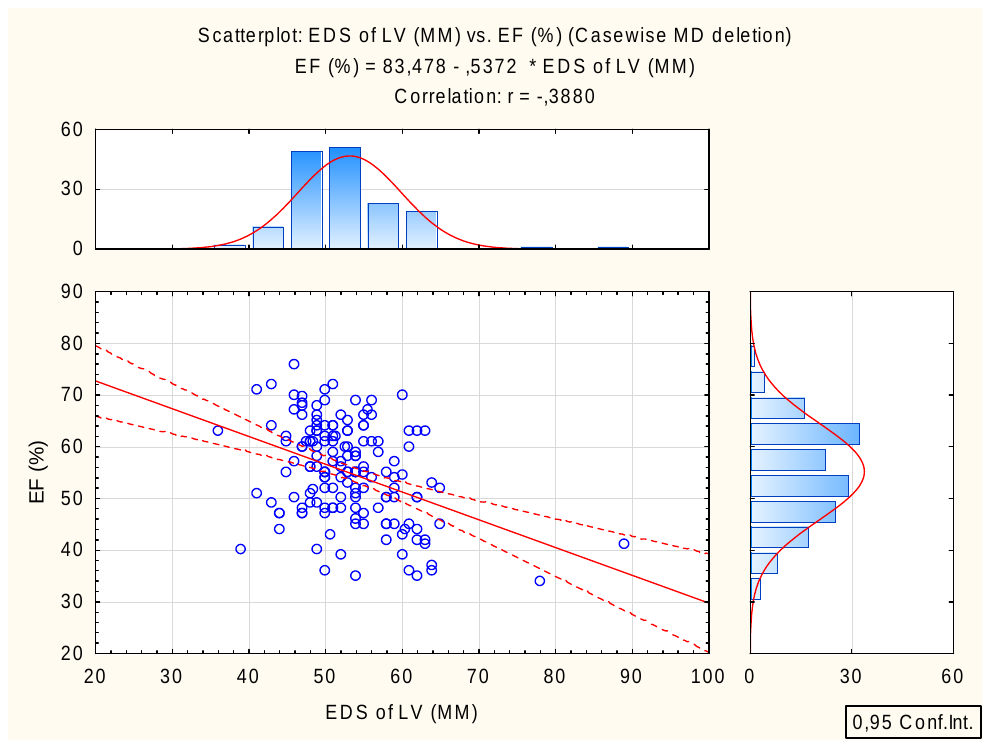

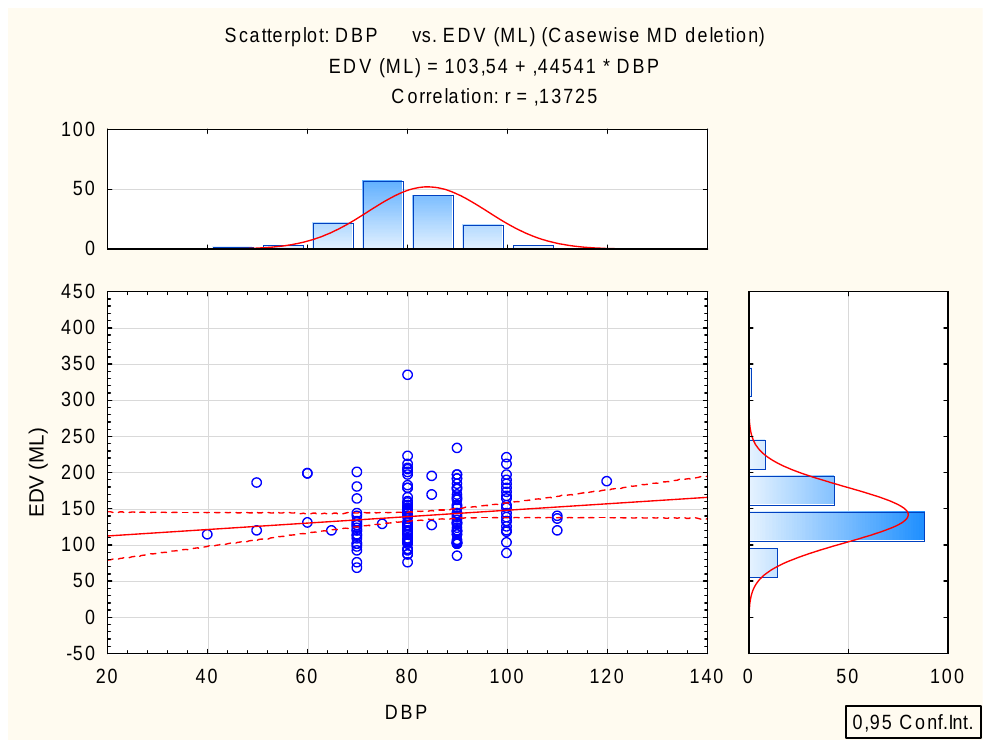

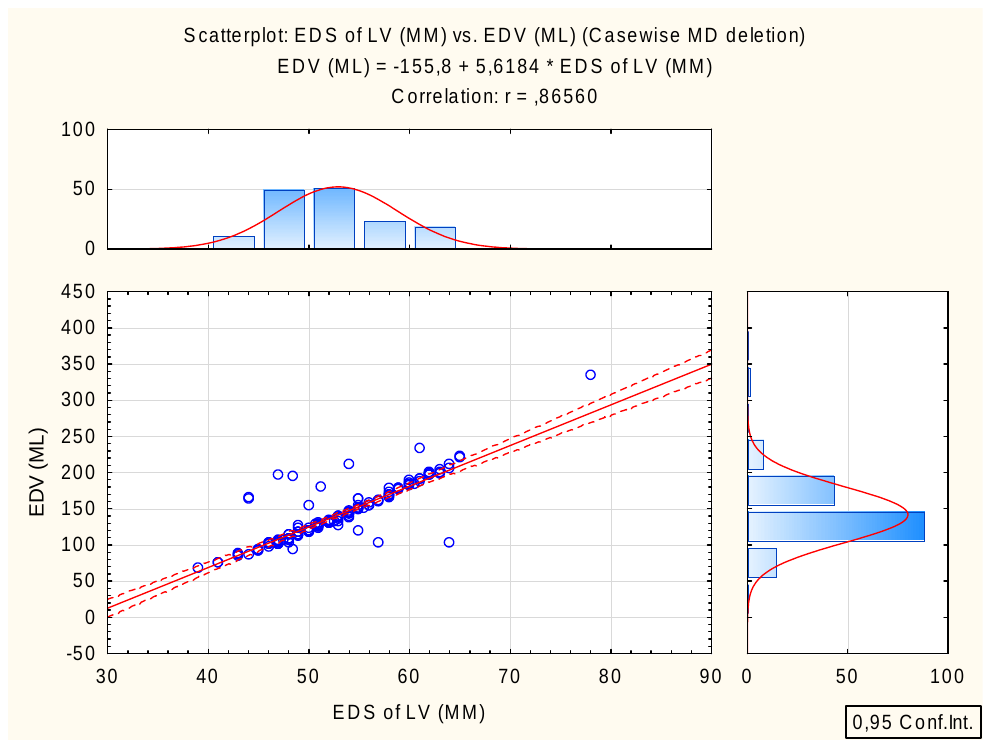

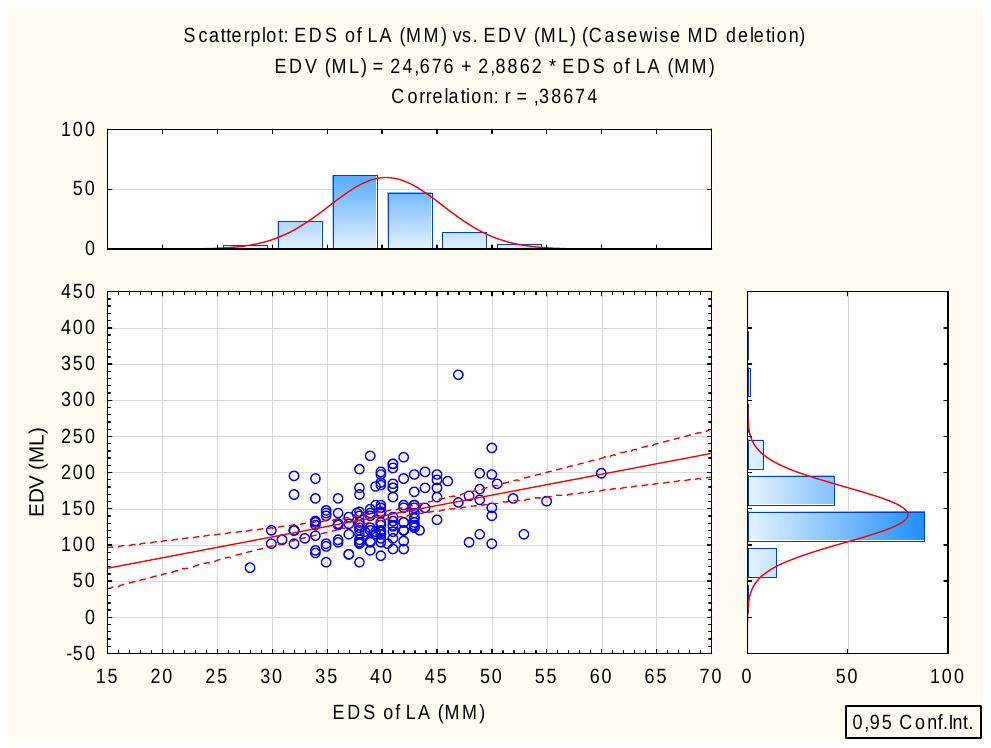

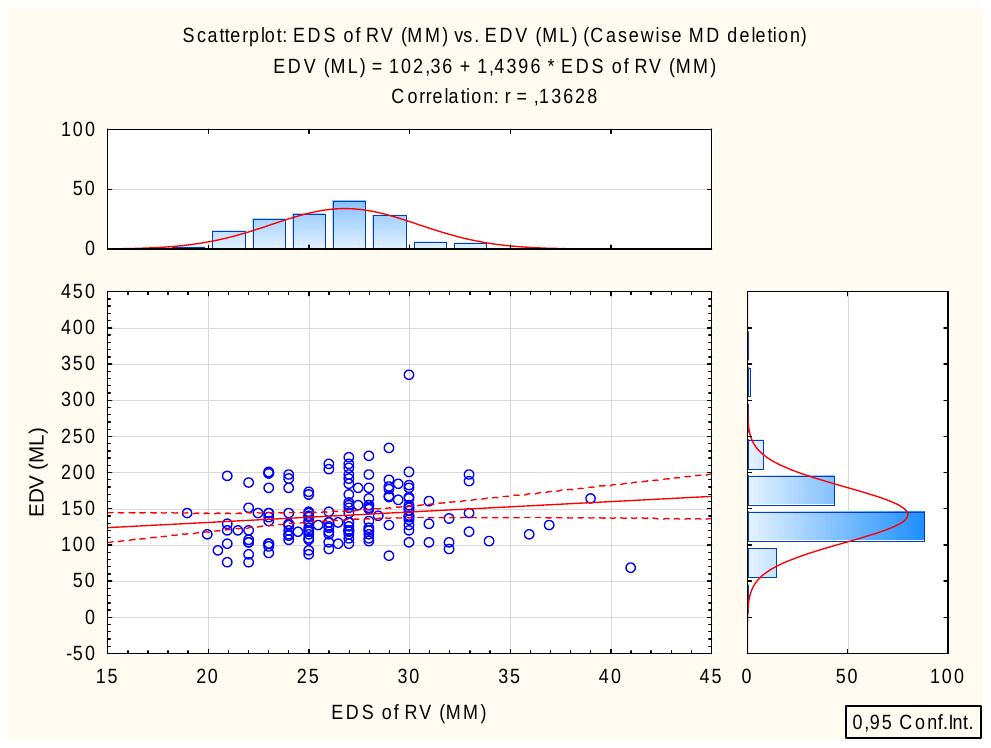

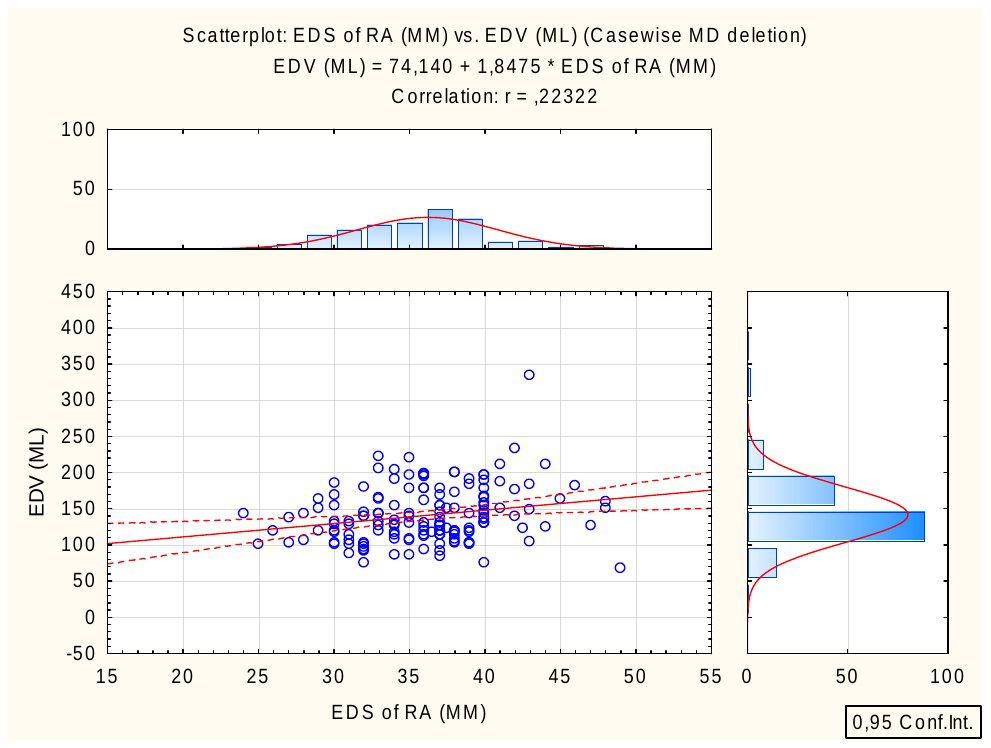

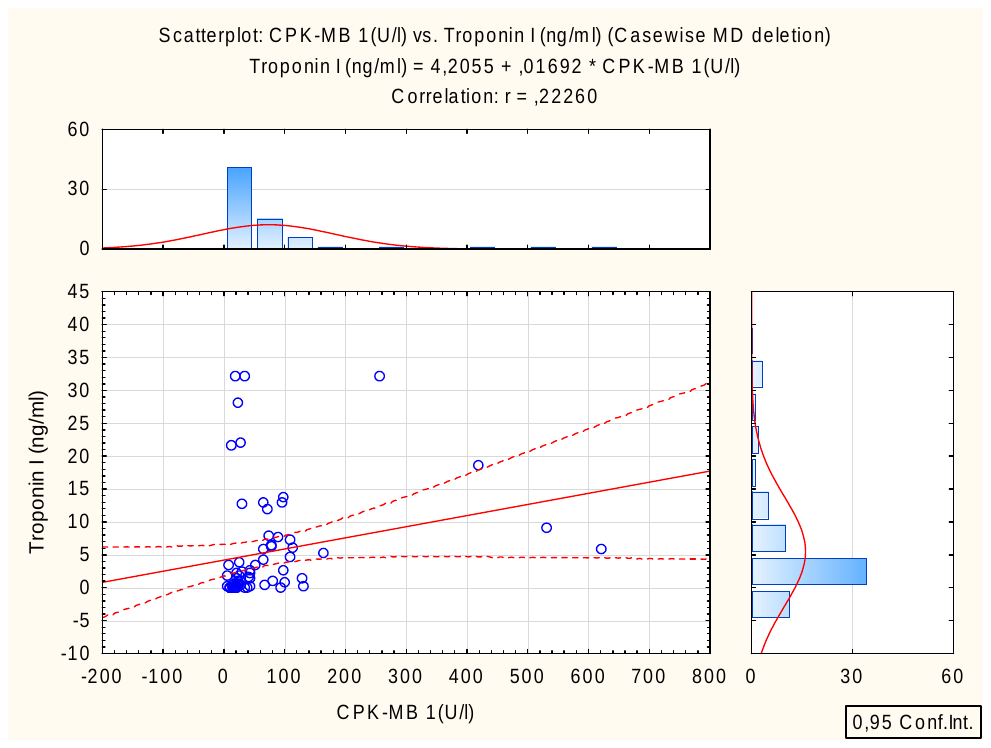

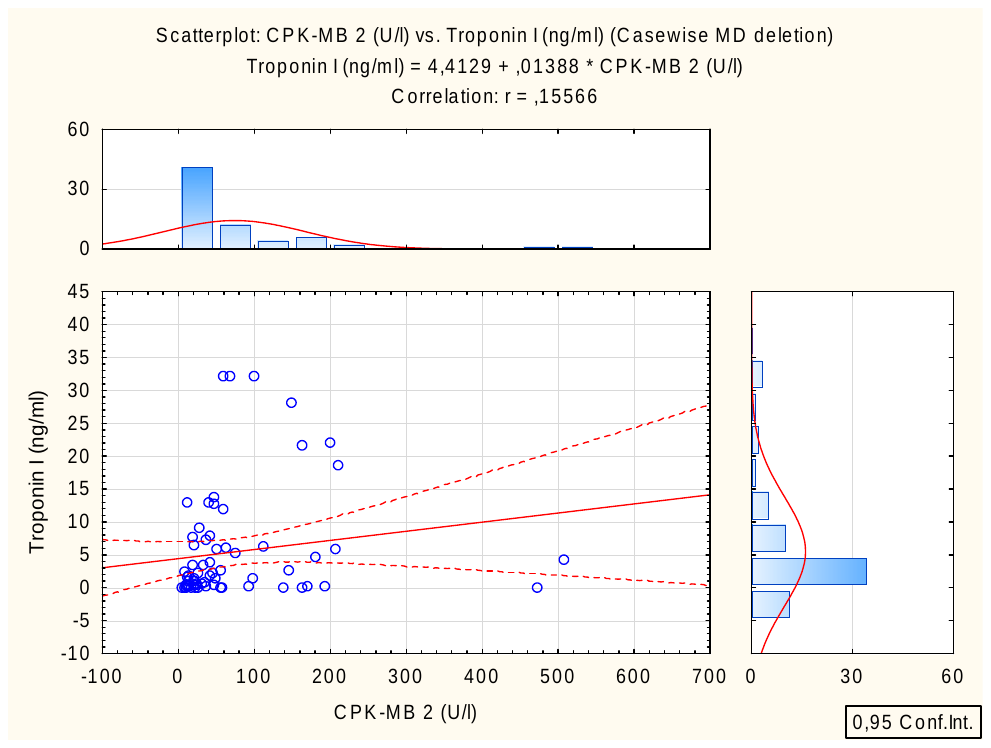

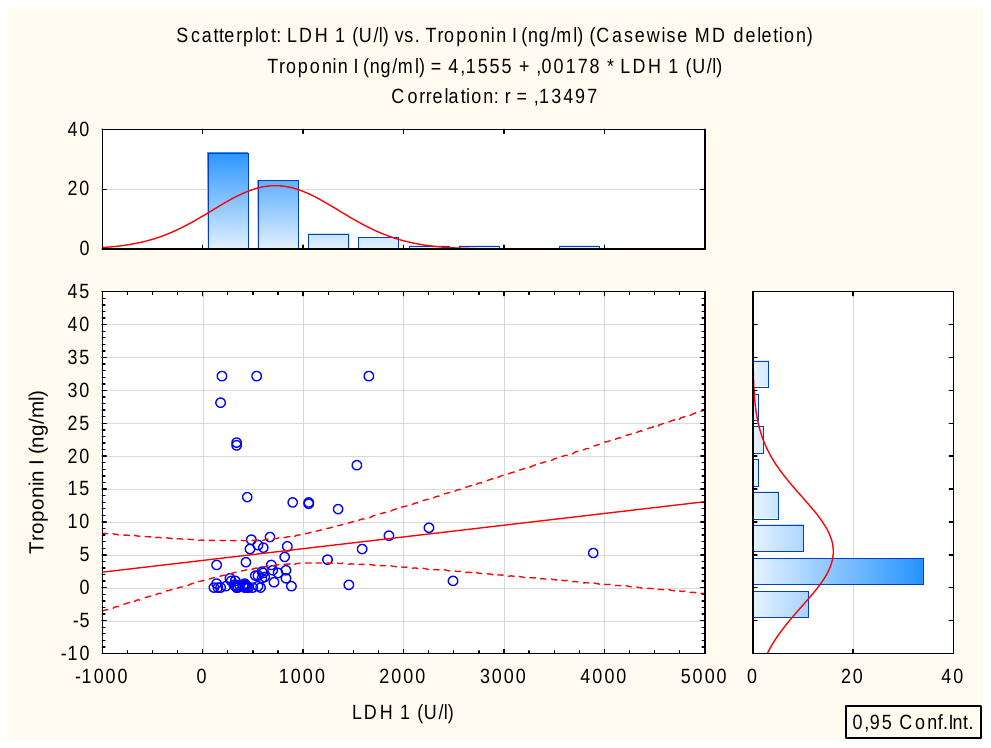

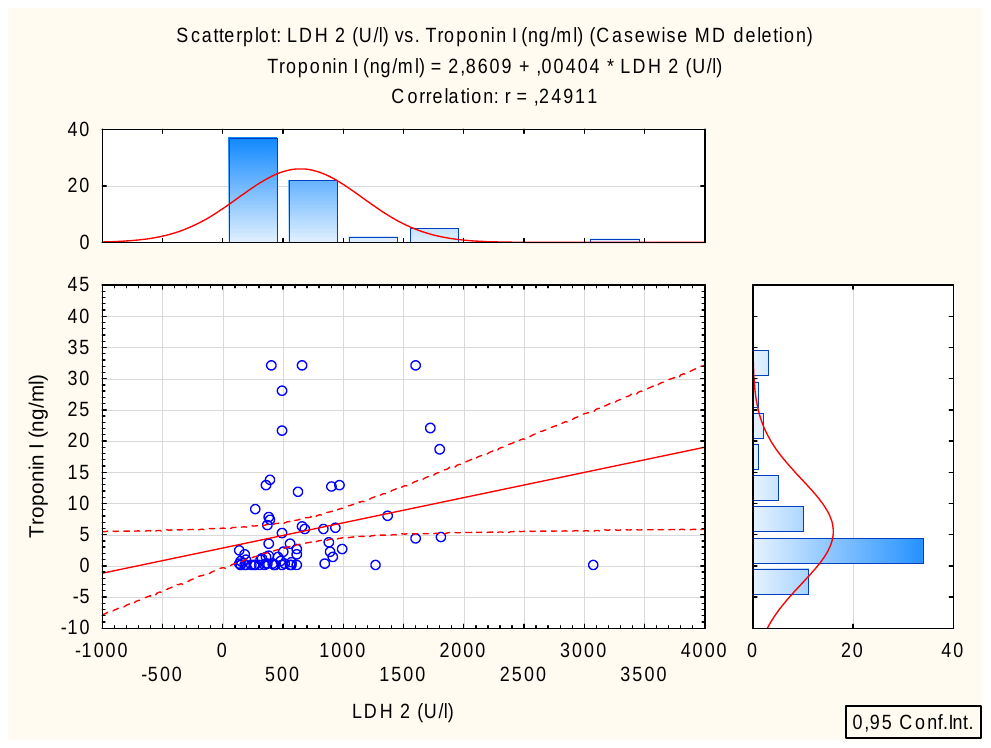

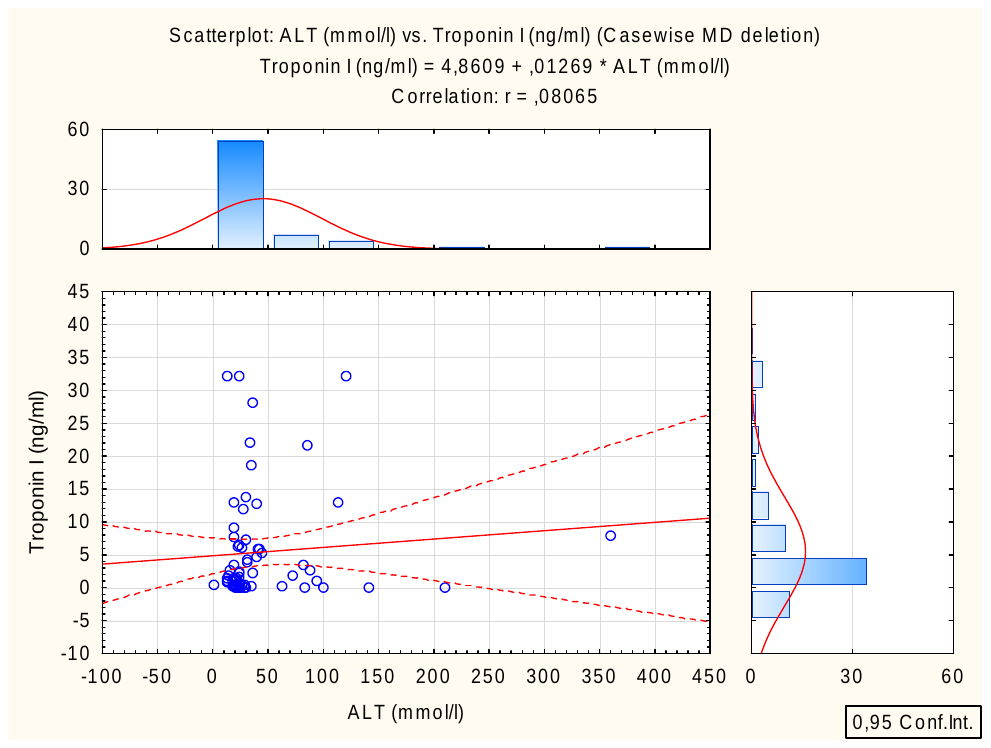

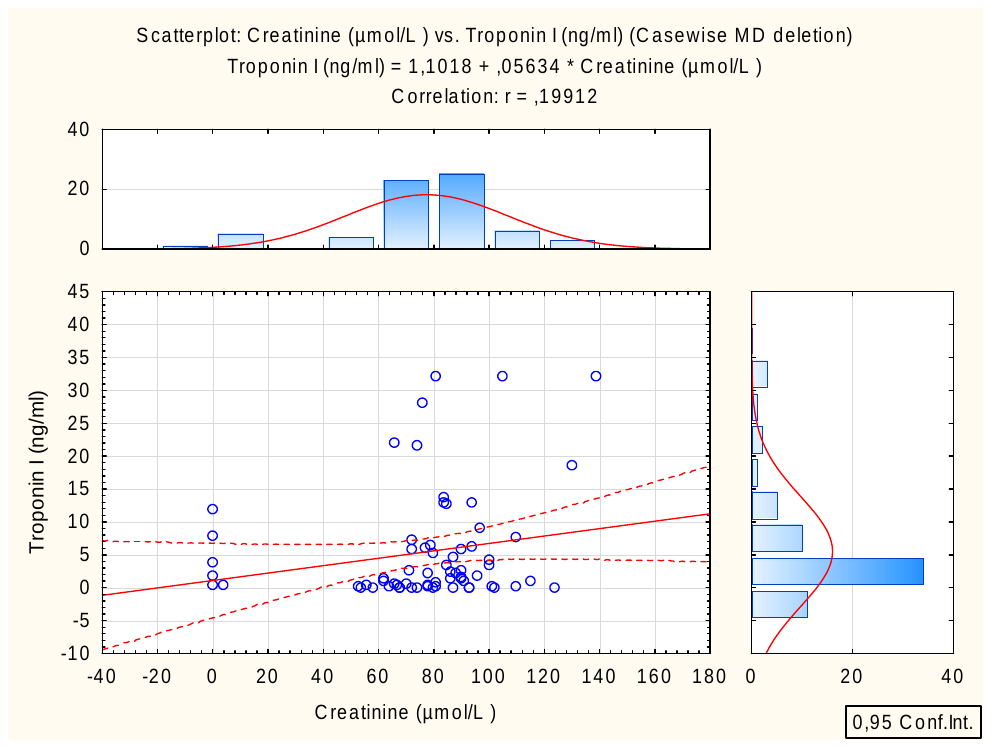
Number of valid cases:191

Observed mean = 61,691099, Observed variance = 106,046184

Distribution: Normal

Parameters: Mean = 61,69110, Variance = 106,0462

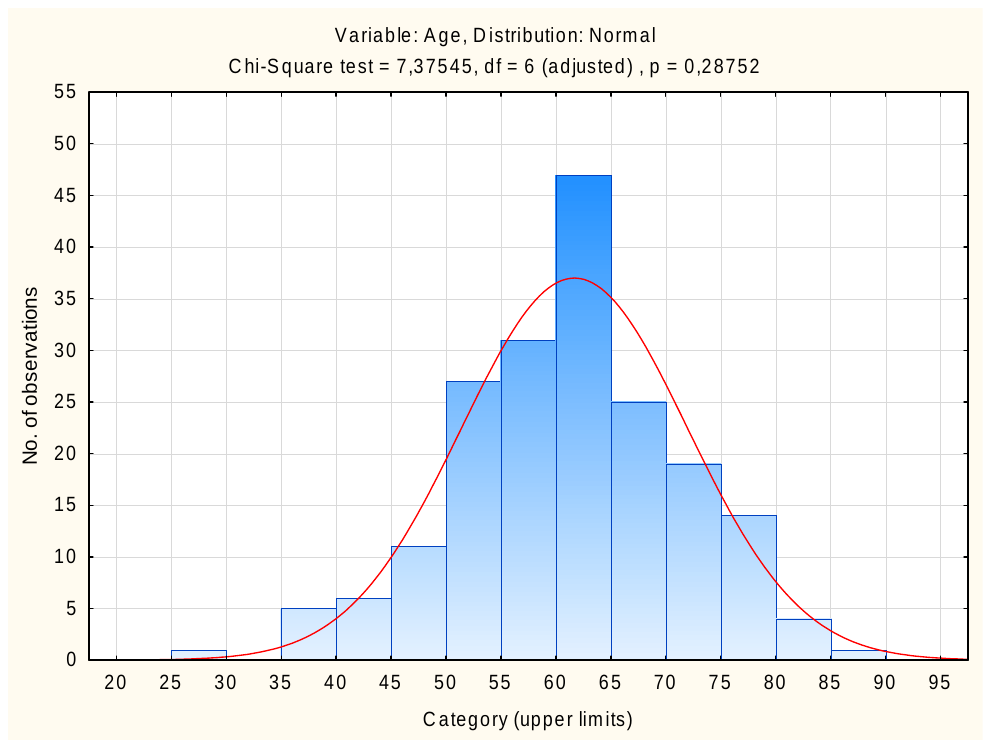
