## Supplementary material for "Early Prognostic Instrumental and Laboratory Biomarkers in Post-MI": Supp. 3

**Supplementary file 3:**

A ROC curve is considered significant if its AUC is significantly different from 0.5.

1) A ROC curve with an AUC of 0.95 or higher would be considered very strong.
2) A ROC curve with an AUC of 0.80 to 0.95 would be considered strong.
3) A ROC curve with an AUC of 0.70 to 0.80 would be considered moderate.
4) A ROC curve with an AUC of 0.60 to 0.70 would be considered weak.
5) A ROC curve with an AUC of less than 0.60 would be considered poor.

It is important to note that the AUC is not the only measure of the performance of a classifier. Other factors, such as sensitivity, specificity, and the clinical context, should also be considered when evaluating a classifier.

1)
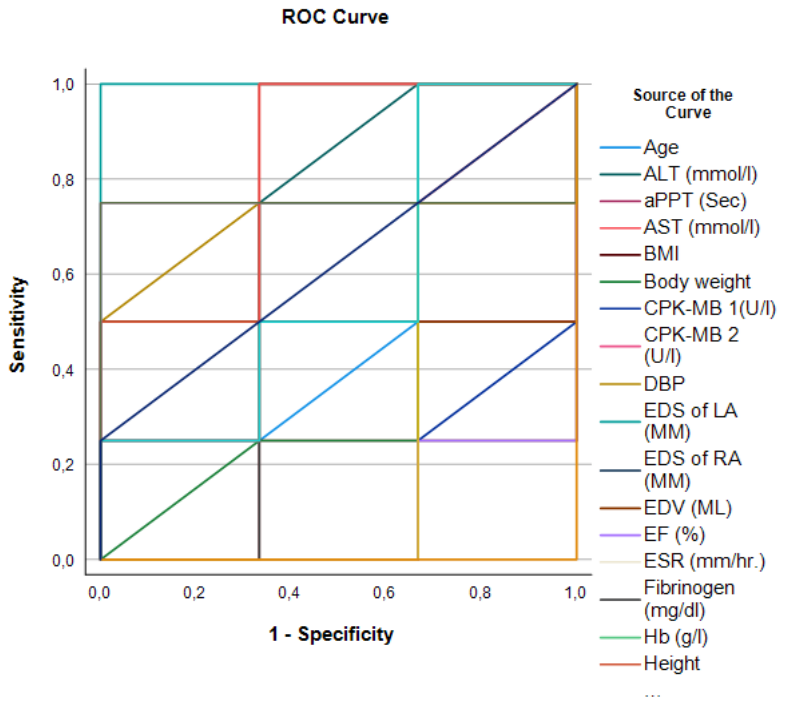
ROC analysis for the arrythmia (Yes) as a complication

| **Area Under the ROC Curve** | |
| --- | --- |
| **Test Result Variable(s)** | **Area** |
| Height | ,833 |
| BMI | ,167 |
| aPPT (Sec) | ,167 |
| CPK-MB 1(U/l) | ,125 |
| Fibrinogen (mg/dl) | ,417 |
| AST (mmol/l) | ,292 |
| LDH 1 (U/l) | ,250 |
| ALT (mmol/l) | ,708 |
| LDH 2 (U/l) | ,750 |
| CPK-MB 2 (U/l) | ,583 |
| ESR (mm/hr.) | ,750 |
| Urine protein | ,625 |
| HR | ,750 |
| Thrombocytes | ,000 |
| Hb (g/l) | ,667 |
| Lymphocytes % | ,583 |
| EF (%) | ,083 |
| EDS of RA (MM) | ,667 |
| EDS of LA (MM) | 1,000 |
| EDV (ML) | ,500 |
| DBP | ,708 |
| Body weight | ,375 |
| Age | ,458 |
| SBP | ,750 |
| The test result variable(s): CPK-MB 1(U/l), AST (mmol/l), ALT (mmol/l), Urine protein, DBP, Body weight, Age has at least one tie between the positive actual state group and the negative actual state group. Statistics may be biased. | |

2) ROC analysis for the concomitant (Yes) disease variable

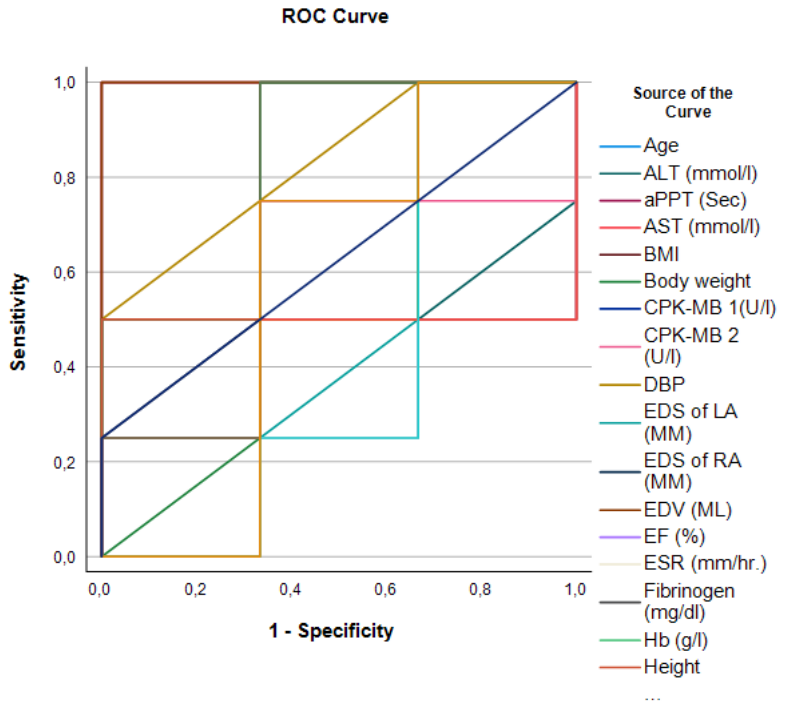

| **Area Under the ROC Curve** | |
| --- | --- |
| **Test Result Variable(s)** | **Area** |
| Age | ,500 |
| HR | ,667 |
| SBP | ,750 |
| DBP | ,833 |
| EF (%) | ,250 |
| EDV (ML) | 1,000 |
| EDS of LA (MM) | ,542 |
| EDS of RA (MM) | ,667 |
| Hb (g/l) | ,833 |
| Lymphocytes % | ,417 |
| Thrombocytes | ,583 |
| ESR (mm/hr.) | ,333 |
| Urine protein | ,625 |
| CPK-MB 1(U/l) | ,500 |
| LDH 1 (U/l) | ,500 |
| CPK-MB 2 (U/l) | ,583 |
| LDH 2 (U/l) | ,417 |
| aPPT (Sec) | 1,000 |
| Fibrinogen (mg/dl) | ,792 |
| AST (mmol/l) | ,333 |
| ALT (mmol/l) | ,375 |
| Height | ,500 |
| BMI | ,583 |
| Body weight | ,458 |
| The test result variable(s): DBP, EDS of LA (MM), Urine protein, Fibrinogen (mg/dl), ALT (mmol/l), Body weight has at least one tie between the positive actual state group and the negative actual state group. Statistics may be biased. | |

3) ROC analysis for the Hypertension (Yes) as a concomitant disease

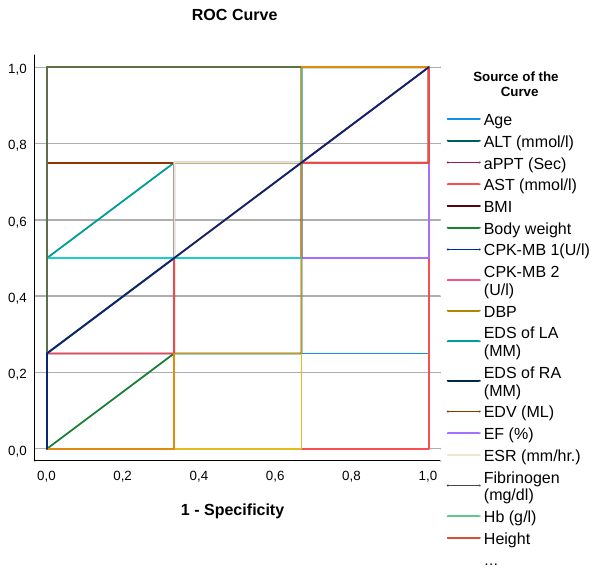

| **Area Under the ROC Curve** | |
| --- | --- |
| **Test Result Variable(s)** | **Area** |
| Age | ,250 |
| HR | 1,000 |
| SBP | 1,000 |
| DBP | 1,000 |
| EF (%) | ,167 |
| EDV (ML) | ,833 |
| EDS of LA (MM) | ,792 |
| EDS of RA (MM) | ,750 |
| Hb (g/l) | 1,000 |
| Lymphocytes % | ,667 |
| Thrombocytes | ,417 |
| ESR (mm/hr.) | ,625 |
| Urine protein | ,625 |
| CPK-MB 1(U/l) | ,333 |
| LDH 1 (U/l) | ,333 |
| CPK-MB 2 (U/l) | ,667 |
| LDH 2 (U/l) | ,500 |
| aPPT (Sec) | ,625 |
| Fibrinogen (mg/dl) | ,542 |
| AST (mmol/l) | ,000 |
| ALT (mmol/l) | ,583 |
| Height | ,625 |
| BMI | ,500 |
| Body weight | ,458 |
| The test result variable(s): EDS of LA (MM), ESR (mm/hr.), Urine protein, aPPT (Sec), Fibrinogen (mg/dl), Height, Body weight has at least one tie between the positive actual state group and the negative actual state group. Statistics may be biased. | |
